# *HLA* autoimmune risk alleles restrict the hypervariable region of T cell receptors

**DOI:** 10.1101/2020.11.08.20227983

**Authors:** Kazuyoshi Ishigaki, Kaitlyn Lagattuta, Yang Luo, Eddie James, Jane Buckner, Soumya Raychaudhuri

## Abstract

Polymorphisms in the *human leukocyte antigen* (*HLA*) genes within the *major histocompatibility complex* (*MHC*) locus strongly influence autoimmune disease risk^1–5^. Two non-exclusive hypotheses exist about the pathogenic role of *HLA* alleles; i) the central hypothesis, where *HLA* risk alleles influence thymic selection so that the probability of T cell receptors (TCRs) reactive to pathogenic antigens is increased^6–8^; and ii) the peripheral hypothesis, where *HLA* risk alleles increase the affinity for pathogenic antigens^9–11^. The peripheral hypothesis has been the main research focus in autoimmunity, while human data on the central hypothesis are lacking. Here, we investigated the influence of *HLA* alleles on TCR composition at the highly diverse complementarity determining region 3 (CDR3), where TCR recognizes antigens. We demonstrated unexpectedly powerful *HLA*-CDR3 associations. The strongest association was found at *HLA-DRB1* amino acid position 13 (n = 628 subjects, explained variance = 9.4%; *P* = 4.1 x 10^−138^). This HLA position mediates genetic risk for multiple autoimmune diseases. In structural analysis of TCR-peptide-MHC complexes, we observed that HLA-DRB1 position 13 does not interact directly with CDR3, but is proximate to antigenic peptide residues that are also close to CDR3. We identified multiple CDR3 amino acid features enriched by *HLA* risk alleles; for example, the risk alleles of rheumatoid arthritis, type 1 diabetes, and celiac disease all increase the hydrophobicity of CDR3 position 109 (*P* < 2.1 x 10^−5^). In the setting of celiac disease, the CDR3 features favored by *HLA* risk alleles are more enriched among candidate pathogenic TCRs than control TCRs (*P* = 2.4 × 10^−6^ for gliadin specific TCRs). Together, these results provide novel genetic evidence supporting the central hypothesis.

## Main

Autoimmune diseases include a broad set of disorders where the immune system attacks self-antigens. For all autoimmune diseases, the *major histocompatibility complex* (*MHC*) locus, harboring the human leukocyte antigen (*HLA*) genes, accounts for more risk than any other locus in the genome^1–5^. For example, 12.7% of the phenotypic variance of rheumatoid arthritis (RA), a prototypical autoimmune disease, can be explained by five amino acid polymorphisms within the *MHC* locus that encode the antigen binding pocket of HLA^1^. In sharp contrast, all established non-*MHC* risk alleles in aggregate explains 4% of the phenotypic variance of RA^12^. Antigenic peptides presented by HLA proteins are recognized by T cell receptors (TCRs), which initiate antigen-specific immune responses. Defining the mechanisms by which *HLA* risk alleles influence the risk of autoimmune diseases remains a critical question.

The commonly referenced explanation for autoimmune risk in the *MHC* locus is that the protein products of risk *HLA* alleles efficiently present critical autoantigens in peripheral lymphoid organs (“peripheral hypothesis” in **Figure 1**)^9–11^. For example, citrullinated self-peptides have higher binding affinity to HLA proteins encoded by *HLA-DRB1* RA risk alleles than those encoded by protective alleles^13,14^. Similar findings have been reported for type 1 diabetes (T1D)^15^ and celiac disease (CD)^16–18^. However, there is an alternative, non-mutually exclusive hypothesis: *HLA* risk alleles may modulate the risk of autoimmunity by influencing thymic T cell selection, resulting in an increased frequency of autoreactive TCRs (“central hypothesis” in **Figure 1**)^6–8^. The TCR antigen specificity is defined by hypervariable complementary determining region 3 (CDR3). During T cell development in the thymus, a highly diverse CDR3 repertoire is generated through random VDJ recombination in immature T cells^19^. Thymic epithelial cells present a family of self-antigenic peptides on HLA proteins. T cells that cannot generate substantial TCR signaling from any HLA-peptide complex die by neglect (positive selection). However, to protect against autoimmunity, the T cell dies by apoptosis if TCR signaling from any complex is too strong (negative selection)^20–22^. Although multiple studies demonstrated the importance of thymic T cell selection in autoimmunity^23–25^, the potential role of the *HLA* risk alleles in shaping the T cell repertoire during thymic selection has yet to be demonstrated in humans.

**Figure 1.**
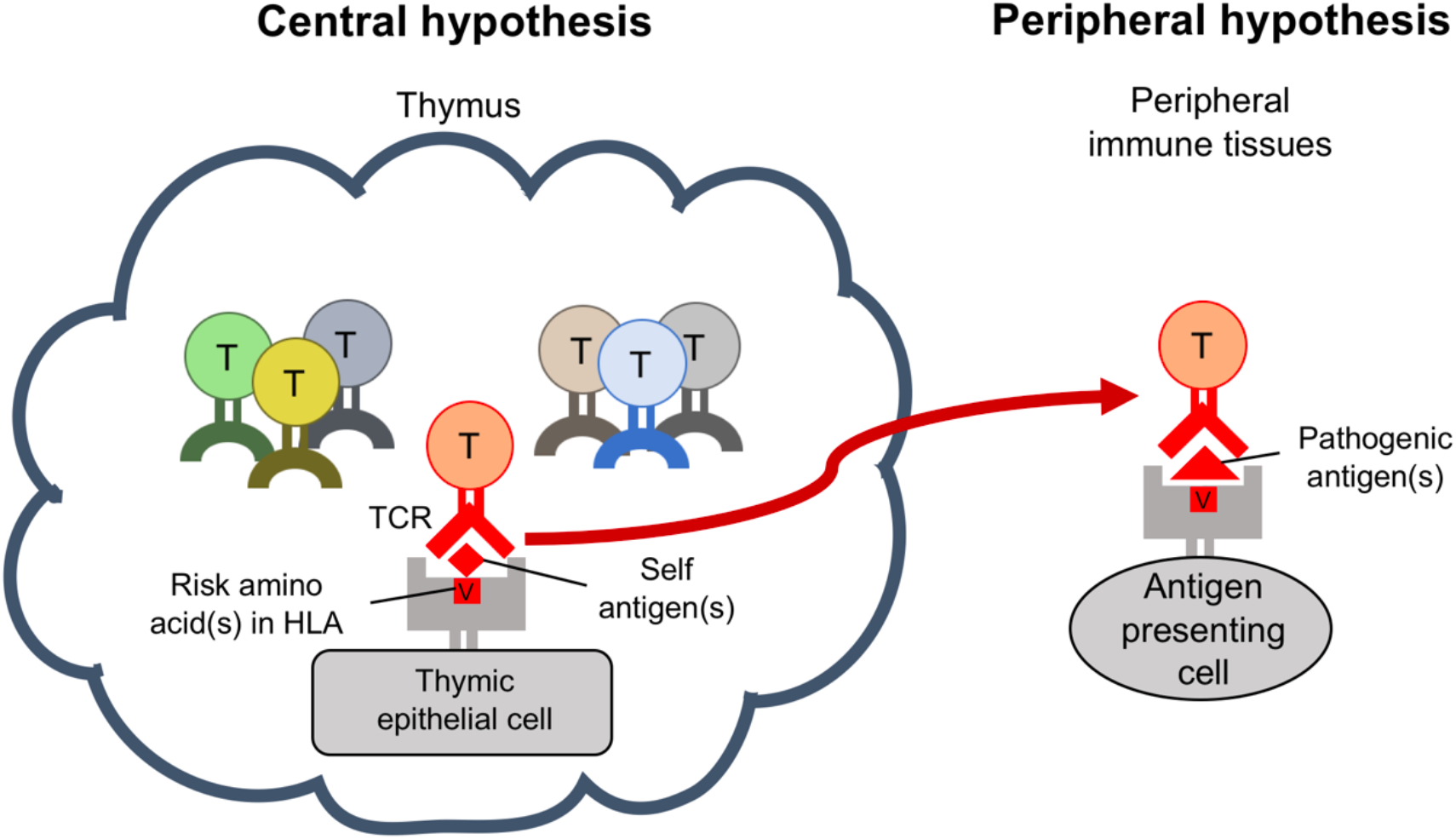
Central vs peripheral hypothesis of *HLA* risk alleles. Pathogenic roles of HLA in autoimmune disease can be explained by two major hypotheses; the central hypothesis (left) and the peripheral hypothesis (right). During T cell development in the thymus, a highly diverse CDR3 repertoire is generated (T cells undergoing selection illustrated in multiple colors). In the central hypothesis, HLA proteins encoded by risk alleles allow more autoreactive TCRs (in red) to survive thymic selection. In the peripheral hypothesis, HLA proteins encoded by risk alleles have higher affinity to critical autoantigens, and therefore are able to more efficiently induce autoimmunity.

Here we sought to assess whether there is genetic evidence supporting the central hypothesis. We treated the amino acid composition of CDR3 as a quantitative trait, and tested its association with *HLA* genotype; we call this CDR3 quantitative trait loci analysis (cdr3-QTL). We then investigated how *HLA* risk alleles modify amino acid compositions of the CDR3 repertoire. Finally, we assessed if the CDR3 features favored by *HLA* risk alleles are enriched in candidate pathogenic TCRs. Our novel TCR analysis framework extends our understanding of HLA-mediated autoimmune risk and provides novel genetic evidence for the central hypothesis.

### Defining CDR3 quantitative traits

We analyzed a public dataset of deeply-sequenced TCR from peripheral blood (n = 628 healthy individuals); we refer to this dataset as the discovery dataset^26^. On average, this dataset has 242,461 unique CDR3 beta chain sequences per individual (**Table 1**). We treated each clone (unique TCR sequence) as a single event (irrespective of read depth). Among unique TCR sequences, 17.9% harbor a frameshift or stop codon; we refer to these sequences as “non-productive” sequences. For almost all analyses, we include only productive sequences to focus on biologically functional TCRs. One important nuance in our analysis is that CDR3 has variable length; CDR3 with 15 amino acids (L15-CDR3) is the most frequent (**Extended Data Figure 1**).

**Table 1.** Characteristics of datasets utilized in this study.

|  |  | **Discovery dataset**<br>**(628 healthy donors)** | **Replication dataset**<br>**(169 healthy donors)** |
| --- | --- | --- | --- |
| **Reference** |  | Emerson et al<sup>26</sup> | Chen L et al<sup>30</sup> |
| **Sample** |  | Peripheral blood | Naïve CD4<sup>+</sup> T cells |
| **TCR data** |  |  |  |
| Sequencing method |  | TCR target sequencing | RNA-seq |
| TCR chains |  | Beta chain | Alpha and beta chains |
| Material |  | DNA | mRNA |
| N. of unique CDR3 per individual | Mean | 242,461 | 1,883 |
|  | S.D | 102,355 | 689 |
| Productive CDR3 ratio | Mean | 82.1% | 94.4% |
|  | S.D | 2.0% | 0.96% |
| **Genotype data** |  |  |  |
| Genome-wide genotype availability |  | No | Yes |
| HLA genotypes |  | Direct typing | Imputed |
| **Ethnicity** |  |  |  |
| Caucasian (n) |  | 348 | 169 |
| Asian or Pacific Islander (n) |  | 19 | - |
| Black or African American (n) |  | 8 | - |
| Native American or Alaska Native (n) |  | 8 | - |
| Unknown (n) |  | 245 | 0 |
| **Age** |  |  |  |
| Mean |  | 39.5 | 57.1 |
| S.D |  | 14.0 | 11.4 |
| **Gender** |  |  |  |
| Male (n) |  | 324 | 72 |
| Female (n) |  | 282 | 97 |
| Unknown (n) |  | 22 | 0 |
| **CMV infection status** |  |  |  |
| Positive (n) |  | 271 | No data |
| Negative (n) |  | 334 |  |
| Unknown (n) |  | 23 |  |

To understand the sequence patterns in CDR3, we calculated the diversity of amino acids at each position and the mutual information across all pairs of positions. The middle positions (108 to 112) are generated by random recombination in the thymus; unsurprisingly these positions have high diversity and little evidence of pairwise mutual information. In contrast, the flanking positions (104-107 and 113-118) are almost exclusively defined by germline encoded V or J genes; hence these positions have little diversity and higher pair-wise mutual information (**Extended Data Figure 1**).

### Identifying cdr3-QTL loci

We first assessed if *HLA* alleles explain amino acid frequencies at specific CDR3 positions. Since the focus of this study is antigen-reactivity of TCR, we restricted our cdr3-QTL analysis to the ten positions (107-116) of CDR3 that directly contact antigens^27^. We refer to locations in HLA amino acid sequences as “sites” and in CDR3 sequences as “positions”. For each CDR3 position, we created a twenty-dimensional phenotype vector representing the frequency of each amino acid across all CDR3 sequences within each individual. We tested the associations between this vector and an HLA site using a multivariate multiple linear regression model (**Methods** and **Extended Data Figure 2**). We included L12-L18 CDR3 (capturing 94.1% of CDR3 sequences) and conducted a total of 24,430 tests: 349 HLA amino acid sites x 70 CDR3 positions (testing each length-position separately). There were 6,410 significant associations (MANOVA test *P* < 2.0 x 10^−6^ = 0.05/24,430; **Supplementary Table 1**). Of these, 76.8% localized to the highly diverse middle positions (108 to 112); in clear contrast, cis-regulatory variants within the *TCR* locus exclusively affect flanking positions correlated with J genes (positions 113-118; **Supplementary Note, Supplementary Figure 1**, and **Supplementary Table 2** and **3**). Of the significant HLA site associations, 81.1% were in a class II HLA protein (**Figure 2a**).

**Figure 2.**
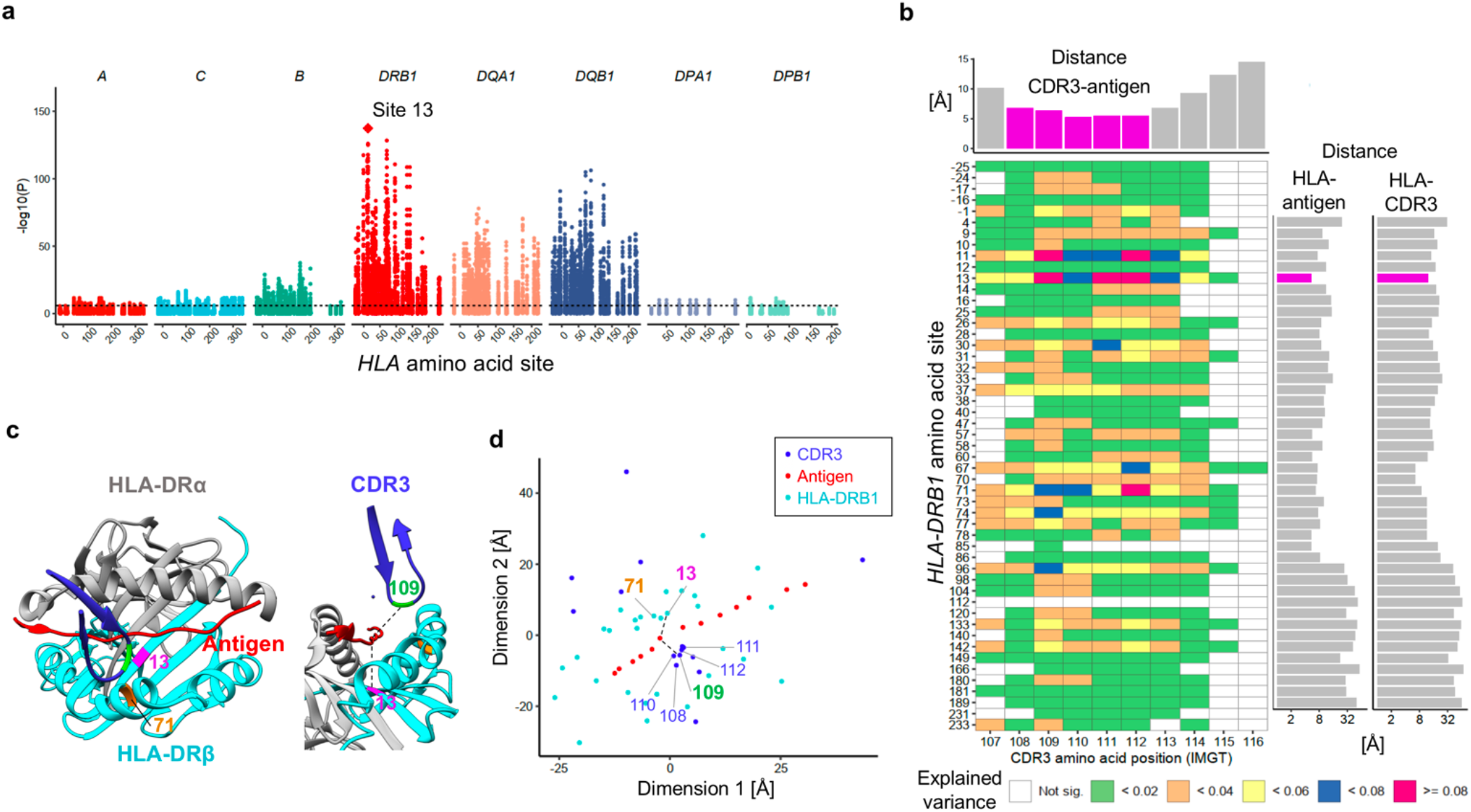
*HLA-DRB1* site 13 strongly influences CDR3 amino acid compositions. **(a)** *P* values in cdr3-QTL analysis using MANOVA (n=628; multivariate multiple linear regression). At each *HLA* site, *P* values of all CDR3 phenotypes are plotted. The *HLA* site with the lowest *P* value (*HLA-DRB1* site 13) is highlighted by a diamond. **(b)** Variance explained in cdr3-QTL analysis (n=628; multivariate multiple linear regression). For each pair of *HLA* site and CDR3 position, the largest variance explained across all CDR3 lengths are depicted in a heatmap. Here, we show all polymorphic sites of *HLA-DRB1* (other *HLA* genes are in **Extended Data Figure 3**). On the top of the heatmap, we present the shortest distances between each CDR3 position and all positions of the antigenic peptide; middle positions of CDR3 were highlighted in magenta. On the side of the heatmap, we also present the shortest distances between each HLA-DRB1 site and all positions of the antigenic peptide (the left bar plot) and those between each HLA-DRB1 site and all positions of CDR3 (the right bar plot); site 13 was highlighted in magenta. Distances were averaged across the five X-ray crystallography structures (**Methods**). **(c)** Structure of HLA-DR, antigenic peptide, and CDR3 protein (Protein database 2IAM). *HLA-DRB1* sites 13, 71 and CDR3 position 109 are highlighted in magenta, orange, and green, respectively. On the left, we depict the antigen (red) and the beta chain CDR3 (dark blue) overlaid onto HLA-DR molecules, looking into the binding groove. On the right, we depict the same complex from a side view. The shortest paths between site 13 and antigenic peptide and those between position 109 to antigenic peptide were shown in black lines. **(d)** Two-dimensional embedding plot based on the pair-wise distances between amino acids of HLA-DRB1, the CDR3 loop, and the antigenic peptide. We down-weighted the distances between HLA and CDR3 so that their antigen-mediated indirect interaction was highlighted (**Methods**). The shortest paths between site 13 and antigenic peptide and those between position 109 to antigenic peptide were shown in black lines as in panel **c**; the paths were well preserved in this embedding.

The strongest association was between HLA-DRB1 site 13 and L13-CDR3 position 109 (MANOVA test *P* = 4.1 x 10^−138^; **Figure 2a-c, Extended Data Figure 3**, and **Supplementary Table 1**). Although CDR3 middle positions are generated by stochastic processes in the thymus, site 13 explained a striking 9.4% of the variance in amino acid usage at position 109. For each CDR3 length, HLA-DRB1 site 13 explained the most variance in amino acid usage (**Supplementary Figure 2**). Even when we controlled the effect of potential confounders (ethnicity, age, sex, cytomegalovirus infection status, and public clonotypes), HLA-DRB1 site 13 always explained the most variance (**Supplementary Figure 3**). Classical alleles were less significantly associated than HLA-DRB1 site 13 (*P* > 4.3 x 10^−96^; **Supplementary Table 4** and **Supplementary Figure 4**). Intriguingly, HLA-DRB1 site 13 is known to drive the risk of multiple autoimmune diseases. This is the residue that explains the most heritability for RA^1,28^ and for juvenile idiopathic arthritis^29^. It represents the strongest association to T1D after HLA-DQB1 site 57^2^.

To reproduce these effects, we obtained a replication data set of 169 healthy individuals consisting of RNA-seq data from sorted naïve CD4^+^ T cells (1,883 unique CDR3 beta chain sequences per individual; **Table1**)^30^. We observed that the variance explained in cdr3-QTL analysis was similar in the replication dataset as in the discovery dataset (Pearson’s r = 0.59), and the greatest variance explained was again identified at *HLA-DRB1* site 13 and L13-CDR3 position 109 (**Supplementary Note, Extended Data Figure 4**, and **Supplementary Table 5**).

To assess if there were independent effects outside of HLA-DRB1 site 13, we conducted serial conditional haplotype analyses within HLA-DRB1 in the discovery data set. In order of descending significance, sites 13, 71, 32, 74, 86 and 30 of HLA-DRB1 showed independently significant signals (**Extended Data Figure 5, 6** and **Supplementary Table 6**). In total, these six sites explained up to 20% of the variation of the middle positions of CDR3, with about half of this variation explained by site 13 (**Extended Data Figure 6**). Among these sites, three (13, 71, and 74) are facing the P4 antigen binding pocket of HLA-DRB1, suggesting that the HLA-DRB1 P4 pocket plays a critical role in shaping the TCR repertoire (**Extended Data Figure 6**). Further conditional analyses outside of *HLA-DRB1* showed signals at both class I and II *HLA* genes; we observed independently significant associations at *HLA-B, HLA-DQB1, HLA-DPB1*, and *HLA-DQA1* with this order of significance (**Extended Data Figure 5**).

To understand if these associations are related to the positioning of residues in the MHC-peptide-TCR complex, we next analyzed five X-ray crystallography-based structures^31–33^. As expected, the antigenic peptide was closer to the middle positions of CDR3, where the cdr3-QTL associations were concentrated, than to flanking positions (6.0 Å vs 14.1 Å on average; one-sided t test *P* = 5.3 x 10^−18^; **Figure 2b, Extended Data Figure 7** and ref^27^). Among all polymorphic sites in *HLA-DRB1*, the site with the most significant cdr3-QTL effect, HLA-DRB1 site 13, was the site closest to the antigenic peptide (5.3 Å on average; **Figure 2b**). In contrast, this site was on average 12.2 Å away from CDR3 residues, a difference which was statistically significant across the five structures examined (one-sided paired t test *P* = 5.6 x 10^−8^; **Figure 2b** and **Extended Data Figure 6**). In addition, all other HLA-DRB1 sites with significant cdr3-QTL effects were also closer to antigenic peptide residues than to CDR3 residues (one-sided paired t test *P* < 0.023; **Extended Data Figure 6** and **Supplementary Figure 5**).

These results suggested that cdr3-QTL signals are mainly driven by indirect interactions between MHC and CDR3 that are mediated by antigenic peptides. To demonstrate this, we calculated pairwise distances between HLA, TCR, and antigen amino acids and embedded them into a two-dimensional space. In this embedding, we preserved distances important for antigen recognition; distances between HLA-DRB1 and antigens and those between TCR and antigens (Pearson’s r > 0.9; **Supplementary Figure 5; Methods**). We observed that in the embedded space, HLA-DRB1 site 13 and CDR3 position 109 were close to a common set of antigen positions, arguing that the association between site 13 and position 109 was mediated by an indirect physical interaction through antigenic peptide residues (**Figure 2d**). The cdr3-QTL analyses so far focused on variance in overall CDR3 composition explained by *HLA*. We next conducted a more detailed analysis to see which specific amino acid pairs co-vary between each HLA site and each CDR3 position (**Methods; Extended Data Figure 2**). Of the 1,262,664 tests that we conducted (923 HLA amino acid alleles x 1,368 CDR3 phenotypes: length-position-amino acid combinations), 15,304 of them were significant (*P* < 3.9 x 10^−8^ = 0.05/1,262,664; **Supplementary Table 7**). A total of 435 of 1,368 CDR3 phenotypes were associated with at least one HLA site amino acid. The effect sizes from the discovery and replication dataset were significantly correlated (Pearson’s r = 0.73; *P* = 1.7 x 10^−71^; **Supplementary Note, Extended Data Figure 4**, and **Supplementary Table 8**). Using this model, we re-evaluated potential impacts of confounders (ethnicity, age, sex, cytomegalovirus infection status, V/J gene usage, and public clonotypes), and confirmed that those effects on cdr3-QTL results were minimal (**Supplementary Figure 6**).

Since we observed consistent cdr3-QTLs between peripheral blood (mixed naïve and memory T cells) and sorted naïve T cells, we hypothesized that cdr3-QTLs might be driven by thymic selection.

There are two alternative possibilities, however: (1) cdr3-QTLs could be driven by genetic biases in V(D)J recombination prior to thymic selection and (2) cdr3-QTLs could be driven by antigen presentation in the periphery subsequent to thymic selection. If (1) is true, then cdr3-QTLs should also be evident in non-productive CDR3s, but we observed no evidence of association for non-productive CDR3s (minimum *P* = 3.7 x 10^−5^ > 0.05/24,430; **Figure 3a-b**). If (2) is true, then T cells with CDR3 sequences favored by specific *HLA* alleles should be clonally expanded; however, cdr3-QTL signals were attenuated when we included clonally expanded TCRs (explained variance reduced by 48.6% on average; **Figure 3c-d**). In addition to the robust replication in naïve CD4^+^ T cells, these results suggest that cdr3-QTLs reflect thymic selection favoring different CDR3 sequence features in the context of different *HLA* alleles (see **Supplementary Note** for more detailed discussion).

**Figure 3.**
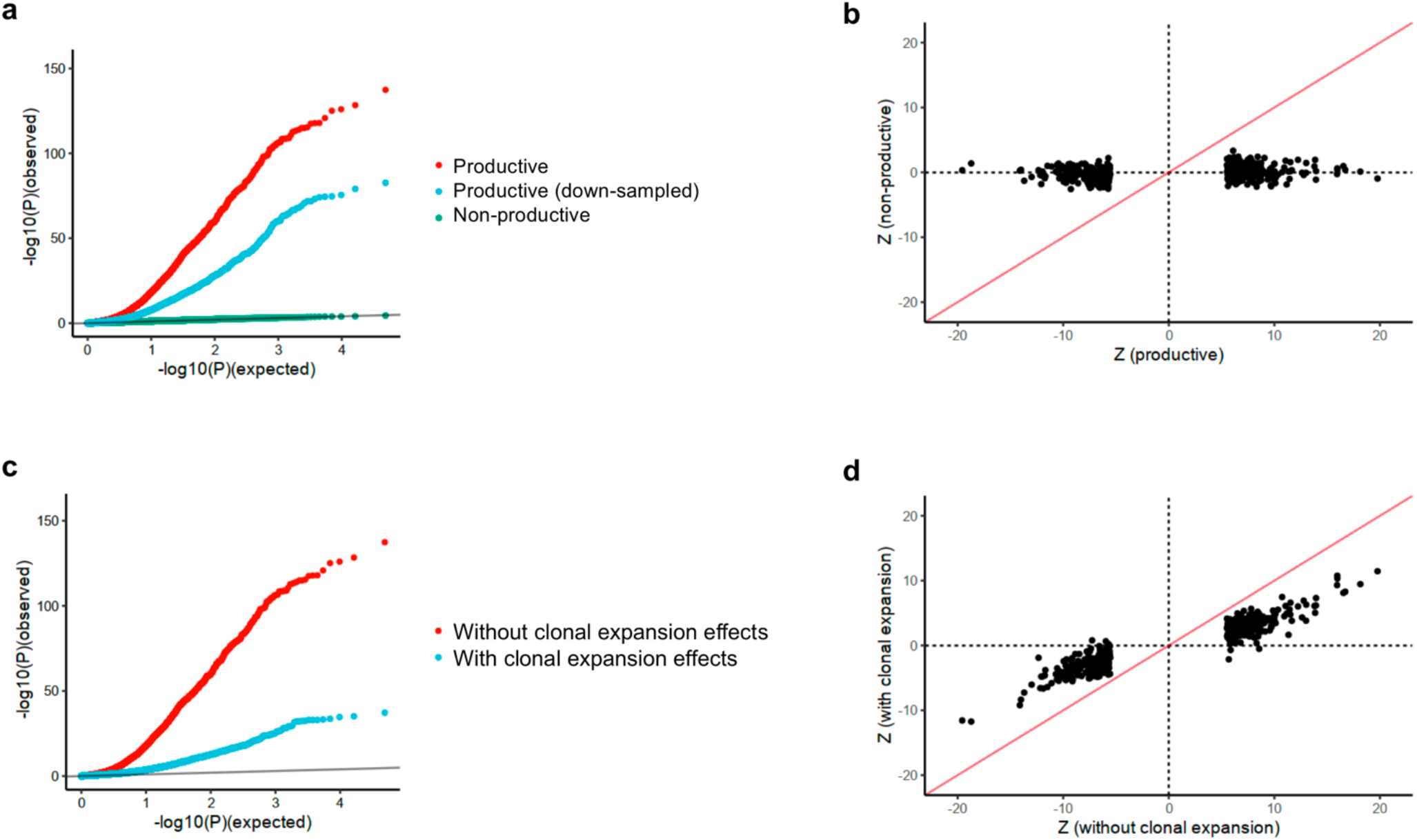
cdf3-QTL effects were not observed for non-productive CDR3 and attenuated by clonal expansion. **(a)** QQ plot of cdr3-QTL analysis in three different settings (n = 628; multivariate multiple linear regression): 1) associations with productive CDR3 sequences (the primary analysis, in red), ii) productive CDR3 sequences down-sampled to match the number of non-productive sequences (in blue), and 3) non-productive CDR3 sequences (in green). **(b)** Z score comparison between cdr3-QTL analyses using productive (the primary analysis) and those using non-productive CDR3 (n = 628; linear regression). **(c)** QQ plot of cdr3-QTL analysis in two different settings (n = 628; multivariate multiple linear regression): 1) considering all CDR3 sequences, including duplicate sequences from clonal expansion (blue) and 2) excluding duplicate CDR3 sequences (the primary analysis, in red). **(d)** Z score comparison between cdr3-QTL analyses with and without duplicate CDR3 sequences from clonal expansion (n = 628; linear regression). The analyses in panel **b** and **d** were restricted to the 435 CDR3 phenotypes (length-position-amino acid combinations) which had at least one significant association in the primary analysis (*P* < 0.05/1,262,664; linear regression).

### Defining and quantifying CDR3 patterns associated with autoimmunity

Since the HLA site that explained the most variance in CDR3 composition (*HLA-DRB1* site 13) was the site with the strongest association to RA risk, we hypothesized that *HLA* risk for RA could be partially mediated by TCR composition. If *HLA* risk for RA is mediated by cdr3-QTLs, the effect sizes of the six possible amino acids at HLA-DRB1 site 13 on RA risk should track with their effects on CDR3 composition. We examined the results at L14-CDR3 position 110, the position among L14-CDR3 for which *HLA-DRB1* site 13 had the strongest signal (MANOVA test *P* = 8.3 x 10^−126^; **Supplementary Table 1**). We observed that site 13 amino acids that raise the risk for RA increase the frequency of aspartic acid (a negatively charged amino acid) while those that protect against RA decrease the frequency of aspartic acid, and that the magnitude of these effects are strongly correlated (Pearson’s r = 0.91; **Extended Data Figure 8**). Similar findings were observed for glutamic acid, another negatively charged amino acid (r = 0.75). Interestingly, we observed the opposite finding for lysine, a positively charged amino acid (r = -0.90; **Extended Data Figure 8**). These results raise the hypothesis that negative charge at position 110 throughout the TCR repertoire is involved in the pathogenesis of RA. Motivated by these results, we aimed to extend our understanding of CDR3 amino acid patterns associated with autoimmune disease risk. Since the risk is driven by multiple *HLA* alleles, analysis of a single *HLA* amino acid site might fail to detect important CDR3 patterns. To directly infer the comprehensive influence of the MHC on CDR3 restriction, we defined a multi-allelic *HLA* risk score for RA, T1D, and CD. Briefly, this *HLA* risk score is a genetic risk score which is a product of two parameters; i) the number of disease-associated *HLA* haplotypes of each individual, and ii) the effect sizes of each haplotype estimated in the previous genetic studies^1,2,5^ (**Figure 4a, Supplementary Table 9** and **Methods**). We calculated *HLA* risk scores in the discovery dataset and assessed which CDR3 patterns are associated using a linear regression model. Out of 1,368 CDR3 phenotypes (combinations of length, positions, and amino acid), we observed significant associations for 94, 206, and 123 phenotypes for RA, T1D, and CD risk scores, respectively (*P* < 3.6 x 10^−5^ = 0.05/1,368; **Supplementary Table 10**). We observed weaker associations to V/J gene usage than to CDR3 patterns (**Supplementary Figure 7**), suggesting that the main target of autoimmune risk is CDR3 compositions, and not V/J genes.

**Figure 4.**
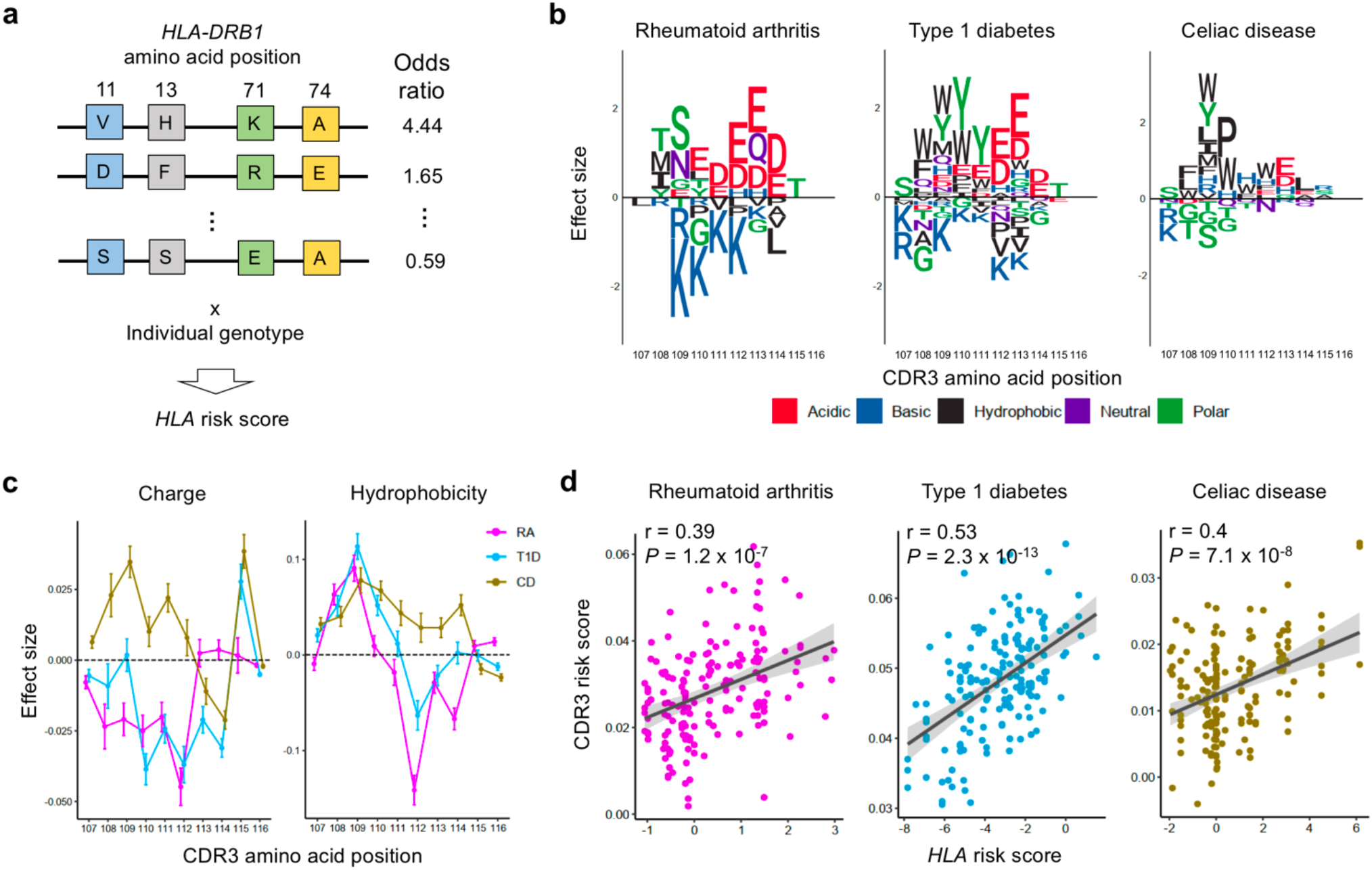
Amino acid patterns influenced by *HLA* risk of RA, T1D, and CD. **(a)** Our strategy to quantify per-individual MHC-wide risk of autoimmune diseases (*HLA* risk score), based on effect size estimates of disease-associated *HLA* haplotypes in previous studies. Haplotypes are depicted as joint amino acid assignments for the polymorphic sites in *HLA-DRB1* (rows). The odds ratio for each haplotype is multiplied by the individual’s dosage (0, 1, 2, if the individual is null, heterozygous, or homozygous, respectively) to compute the *HLA* risk score. **(b)** CDR3 amino acids influenced by *HLA* risk score for RA, T1D and CD. We conducted cdr3-QTL analysis using the *HLA* risk score for each of these diseases separately, testing for differential usage of each amino acid at each position of each length CDR3 (n=628; linear regression). To create a sequence logo for each disease, the effect sizes of significant (*P* < 0.05/1,368) associations for each amino acid at a given position were summed across L12-L18 CDR3s. **(c)** CDR3 amino acid features influenced by *HLA* risk score. We conducted cdr3-QTL analysis using *HLA* risk score for RA, T1D, and CD; the CDR3 phenotypes were amino acid features at each position of each length CDR3 (n=628; linear regression). The effect sizes for each feature at a given position were meta-analyzed using a fixed effect model and the results of two features were provided with S.E (see **Extended Data Figure 10** for other features). **(d)** The correlation between *HLA* risk score and CDR3 risk score in the replication dataset for RA, T1D, and CD (n=169; naïve CD4^+^ T cells). Pearson’s *r* is provided.

**Figure 5.**
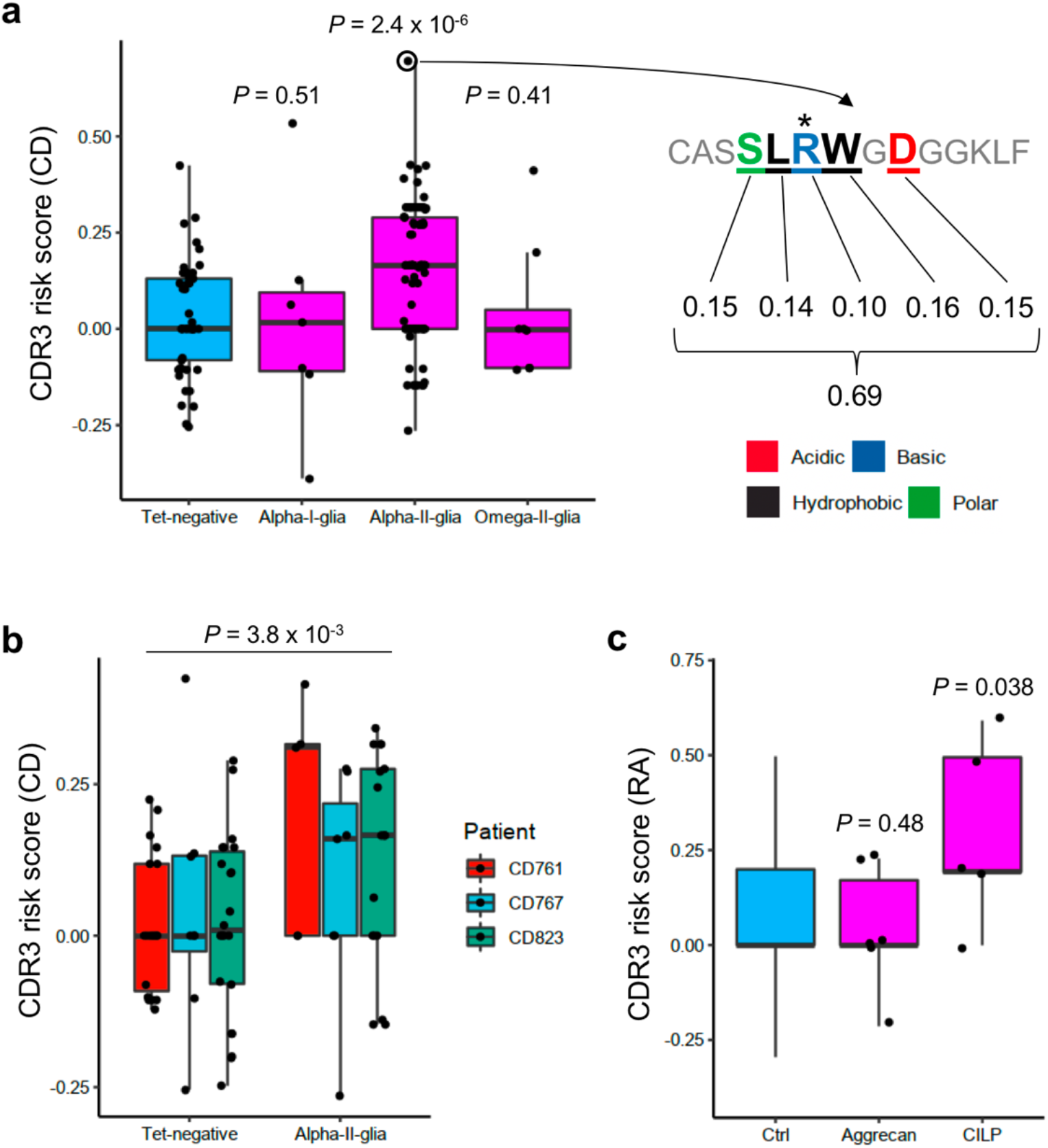
Pathogenic CD4^+^ T cells possess high CDR3 risk score. **(a)** CD-CDR3 risk scores were compared among CD4^+^ T cells with different antigen specificities. Seven TCRs specific to α-I gliadin (n = 4 patients), 92 TCRs specific to α-II gliadin (n = 13 patients), eight TCRs specific to ω-II gliadin (n = 2 patients), and 49 control TCRs (n = 3 patients) were analyzed. An illustration of CDR3 risk score calculation is provided for the CDR3 sequence with the highest score: effect sizes of CDR3 amino acids with a significant association to CD-*HLA* risk score (five amino acids of this sequence) are summed. Arginine (R) at position 109 (*) is known to be important for the recognition of α-II gliadin. One-sided t test *P* values are provided. **(b)** The same analysis as in panel **a** but restricted to three patients from whom we obtained data for both of α-II gliadin specific TCRs (29 sequences) and control TCRs (49 sequences). The *P* value from a linear regression adjusted for donor-level effects is given. **(c)** RA-CDR3 risk scores were compared among CD4^+^ T cells with different antigen specificities. Six TCRs specific to citrullinated aggrecan (n = 5 patients) and five TCRs specific to citrullinated CILP (n = 2 patients) were analyzed. We prepared 1,753 control TCR sequences from an individual homozygous for *HLA-DRB1*\*0401, the *HLA-DRB1* allele with the highest risk for RA (**Methods**). One-sided t test *P* values are provided. Within each boxplot, the horizontal lines reflect the median, the top and bottom of each box reflect the interquartile range (IQR), and the whiskers reflect the maximum and minimum values within each grouping no further than 1.5 x IQR from the hinge.

To illustrate the CDR3 amino acid patterns characteristic of each disease, we created sequence logos (**Methods; Figure 4b;** stratified by CDR3 length, **Extended Data Figure 9**) and noted that amino acids with similar biochemical features demonstrated similar trends. This suggested that there may be latent biochemical features driving these disease-specific cdr3-QTLs. To quantify this, we examined the five broad aggregate amino acid features; charge, hydrophobicity, refractivity, propensity for canonical secondary structures, and molecular size (**Methods**). At each position of each length of CDR3, we calculated the weighted average of each feature to create 350 CDR3 phenotypes in total (5 features x 70 positions; **Methods**). We then tested their association with the *HLA* risk score using a linear regression (**Figure 4c, Extended Data Figure 10**, and **Supplementary Table 11**). Consistent with the above results, RA risk was associated with decreased amino acid charge at multiple positions, including position 110 (*P* < 3.2 x 10^−5^; **Figure 4c**). Interestingly, increased hydrophobicity at position 109 was associated with the *HLA* risk score of all three diseases (*P* < 2.1 x 10^−5^; **Figure 4c**). Stadinksi *et al*. have proposed that elevated hydrophobicity at this position promotes T cell autoreactivity based on experimental work in a type 1 diabetes mouse model^34^. Our results reveal a genetic basis for this CDR3 phenotype and demonstrate its relevance in humans with respect to multiple autoimmune diseases. Moreover, our results suggest that there are several other CDR3 biochemical features associated with T1D, CD, and RA risk individually (**Extended Data Figure 10**).

These CDR3 patterns associated with autoimmune disease risks might indicate T cell reactivity to pathogenic antigens. We developed a scoring system that quantifies the enrichment of these patterns in a given CDR3 sequence (we refer to this as the CDR3 risk score). We utilized CDR3 phenotypes (combinations of lengths, positions, and amino acids) which were significantly associated with the *HLA* risk score in the previous analysis (**Supplementary Table 10**). The CDR3 risk score is the sum of effect sizes when their target phenotype exists in a given CDR3 sequence (**Supplementary Figure 8** and **Methods**). We then evaluated the performance of CDR3 risk score using an independent replication dataset of na’ve CD4^+^ T cells (n = 169). Reassuringly, CDR3 risk scores were significantly correlated with the *HLA* risk scores; Pearson’s r was 0.39, 0.53, and 0.4 for RA, T1D, and CD, respectively (**Figure 4d**). We thus established and validated a method to quantify CDR3 patterns associated with autoimmunity.

### Pathogenic CD4^+^ T cells possess high CDR3 risk scores

Finally, we applied the CDR3 risk score to CD4^+^ T cell populations whose TCRs recognize candidate pathogenic antigens. If the central hypothesis is true, TCRs reactive to pathogenic antigens may have higher CDR3 risk scores. Although our understanding about pathogenic antigens in human autoimmunity is generally limited, promising candidates have emerged for a few autoimmune diseases, and we focused on our analyses on these.

We first analyzed public datasets of TCR sequences from CD patients. The hallmark of CD is the immune reaction to gliadin, a component of gluten, and several different deamidated epitopes have been reported to be pathogenic in CD; α-I, α-II, and ω-II gliadin^35^. From previous studies where HLA-DQ2 tetramers were utilized to select antigen-specific T cell populations, we collected sequences of TCR which are specific to these three epitopes, and control non-specific TCRs^36–38^; seven TCRs specific to α-I gliadin (n = 4 patients), 92 TCRs specific to α-II gliadin (n = 13 patients), eight TCRs specific to ω-II gliadin (n = 2 patients), and 49 control TCRs (n = 3 patients) (**Supplementary Table 12**). Strikingly, we observed that α-II gliadin-specific TCRs have higher CD-CDR3 risk score than control TCRs (one-sided t test *P* = 2.4 x 10^−6^; **Figure 5a**). Interestingly, a CDR3 sequence with the highest score was reactive to α-II gliadin, and it had arginine at position 109, which is known to be important for the recognition of α-II gliadin^38,39^ (**Figure 5a**). In contrast, we did not observe significant differences in CD-CDR3 risk scores between TCRs specific to the other gliadin epitopes and control TCRs (one-sided t test *P* > 0.05; **Figure 5a**); this might indicate that T cell reactivity to α-II gliadin is more relevant for the genetic risk of CD than others. Recognizing that subtle *HLA* genotype differences could affect CDR3 scores, we next conducted an intra-individual analysis by restricting the analysis to the TCR from the three individuals for whom we have control TCR data. Even in this stringent analysis, we still observed significant differences in CD-CDR3 risk score between α-II gliadin-specific and control TCRs (*P* = 3.8 x 10^−3^; **Figure 5b**).

We then analyzed TCR data from RA patients. The hallmark of RA is the immune reaction to citrullinated antigens. We sequenced six TCRs specific to citrullinated aggrecan (n = 5 patients) and five TCRs specific to citrullinated cartilage intermediate layer protein (CILP) (n = 2 patients), which were identified by HLA-DRB1*04:01 or HLA-DRB1*04:04 tetramers. Since we did not have control TCR data from the same individuals, we prepared 1,753 control TCR sequences from an individual homozygous for *HLA-DRB1*\*0401, the allele with the highest *HLA*-risk of RA (**Methods**). TCRs specific to citrullinated CILP have nominally higher RA-CDR3 risk score than control TCRs (one-sided t test *P* = 0.038; **Figure 5c**). However, we did not observe significant differences in the risk score between TCRs specific to citrullinated aggrecan and control TCRs (one-sided t test *P* > 0.05). The inter-epitope differences in CDR3 risk score might be useful to differentiate causal epitopes from those targeted by epitope spreading. However, we need to be cautious about its interpretation considering the limited number of TCRs specific for some antigens. Together, our analyses provide novel genetic evidence supporting that the *HLA* risk increase the frequency of TCRs reactive to, at least some, candidate pathogenic antigens.

## DISCUSSION

Our study demonstrated large effect size associations between MHC polymorphisms and CDR3 amino acid compositions using a novel quantitative trait analysis framework for the TCR. We identified CDR3 amino acid patterns associated with MHC-wide risk for autoimmune diseases which were enriched in T cells reactive to candidate pathogenic antigens. Using the CDR3 risk score, we could prioritize pathogenic T cell populations solely based on TCR sequencing, which might complement tetramer-based analyses.

Although previous studies also investigated covariance between MHC polymorphisms and the TCR repertoire^26,40^, such analyses were restricted to public clonotypes, which represent a small fraction of the repertoire. Associations found with this approach were enriched in the clonally expanded and pathogen-experienced T cell populations.^26,40^ In contrast, our study analyzed the entire repertoire with a novel quantitative framework and consequently captured more comprehensive patterns of HLA-TCR covariance. These associations were attenuated by the inclusion of clonally expanded TCRs, and were robustly replicated in sorted naïve T cells. Thus, while previous methods emphasized HLA-TCR biology in peripheral tissues, our framework highlights HLA-TCR interactions in the thymus.

It is important to recognize that our investigation was limited to the beta chain of the TCR. Though the beta chain is more important for antigen-specificity and therefore the central hypothesis^27^, future work is needed to assess whether there are also disease-relevant cdr3-QTLs affecting the alpha chain of the TCR. Additionally, because CD4^+^ T cells are nearly twice as prevalent as CD8^+^ T cells in peripheral blood, our analysis was better powered to detect class II HLA-CD4^+^ associations. Future studies in sorted CD8^+^ T cells will be necessary to resolve the relative contribution of MHC class I. Our results highlight the importance of high-throughput experimental systems to identify cognate antigens of CD4^+^ T cells. These technologies have the potential to expand our knowledge of pathogenic antigens of human autoimmunity, which will be essential in combination with this data to establish the role of the central hypothesis.

It is now clear that MHC risk polymorphisms modulate the process of thymic selection and give rise to TCR repertoires that may be poised for autoreactivity. This finding presents a paradigm shift for *HLA* pathogenicity in human autoimmunity, reinvigorating a role for the central hypothesis in mediating inter-individual differences in autoimmune disease risk.

## Extended Data Figures

**Extended Data Figure 1.**
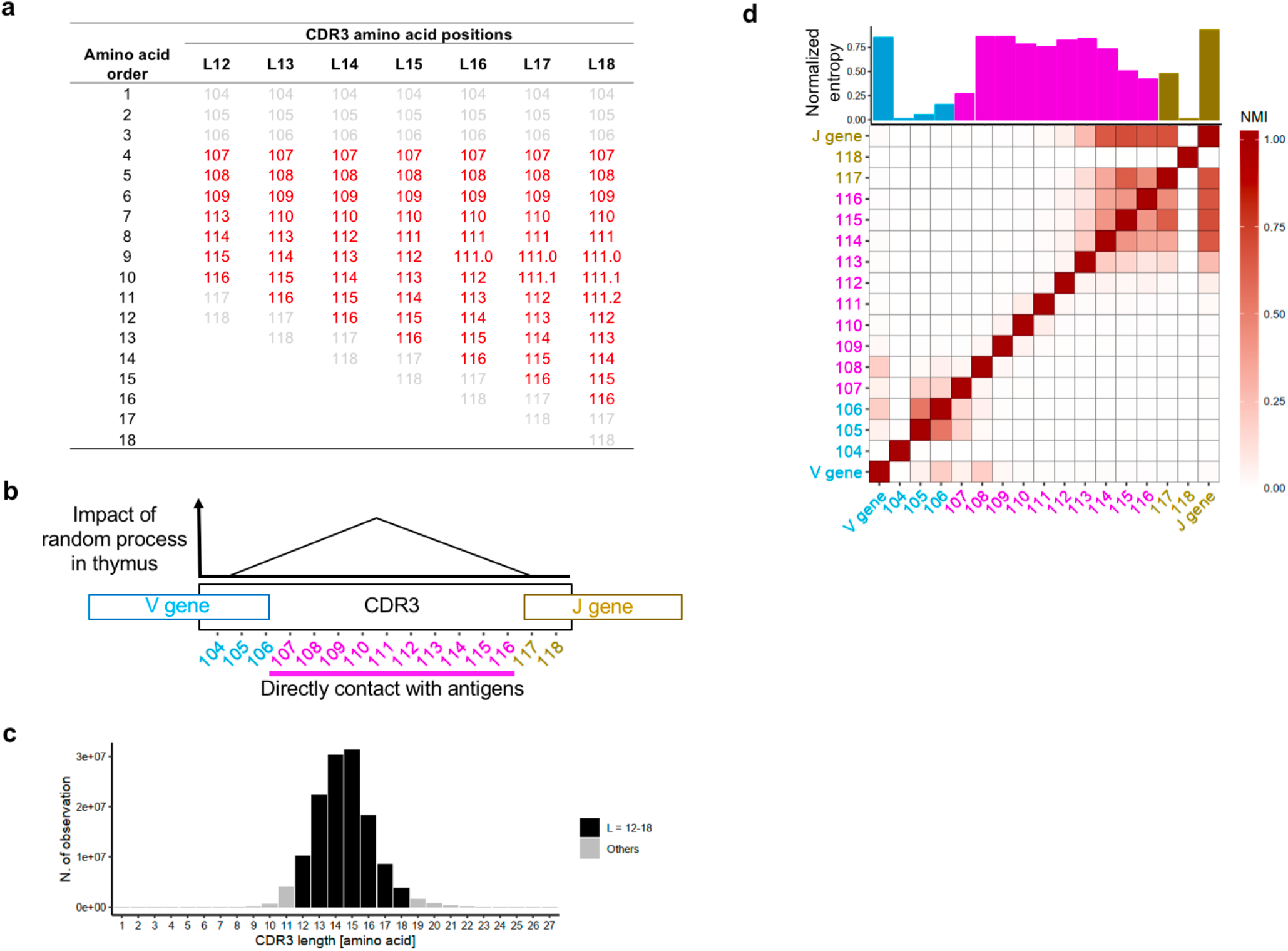
TCR structure in the discovery dataset. **(a)** The amino acid positioning rule in this study. Some middle positions are missing in shorter CDR3s and there are some extra middle positions in longer CDR3s. **(b)** Schematic explanation of the structure of CDR3. During T cell development in thymus, TCR are generated by randomly recombining component genes (V, D, and J gene for beta chain). In addition, several nucleotides are randomly added or deleted at the junctional regions. **(c)** The distribution of CDR3 amino acid length in the discovery dataset. **(d)** The diversity and information of amino acid composition at CDR3 positions (length=15 amino acids). Normalized entropy (bar plot) and normalized mutual information (NMI, heatmap) of amino acid usage at each position of CDR3 and V/J gene usage were calculated in each individual, and the averaged values were provided. CDR3 positions 107-116 which directly contact antigens were highlighted in red (panel **b** and **d**).

**Extended Data Figure 2.**
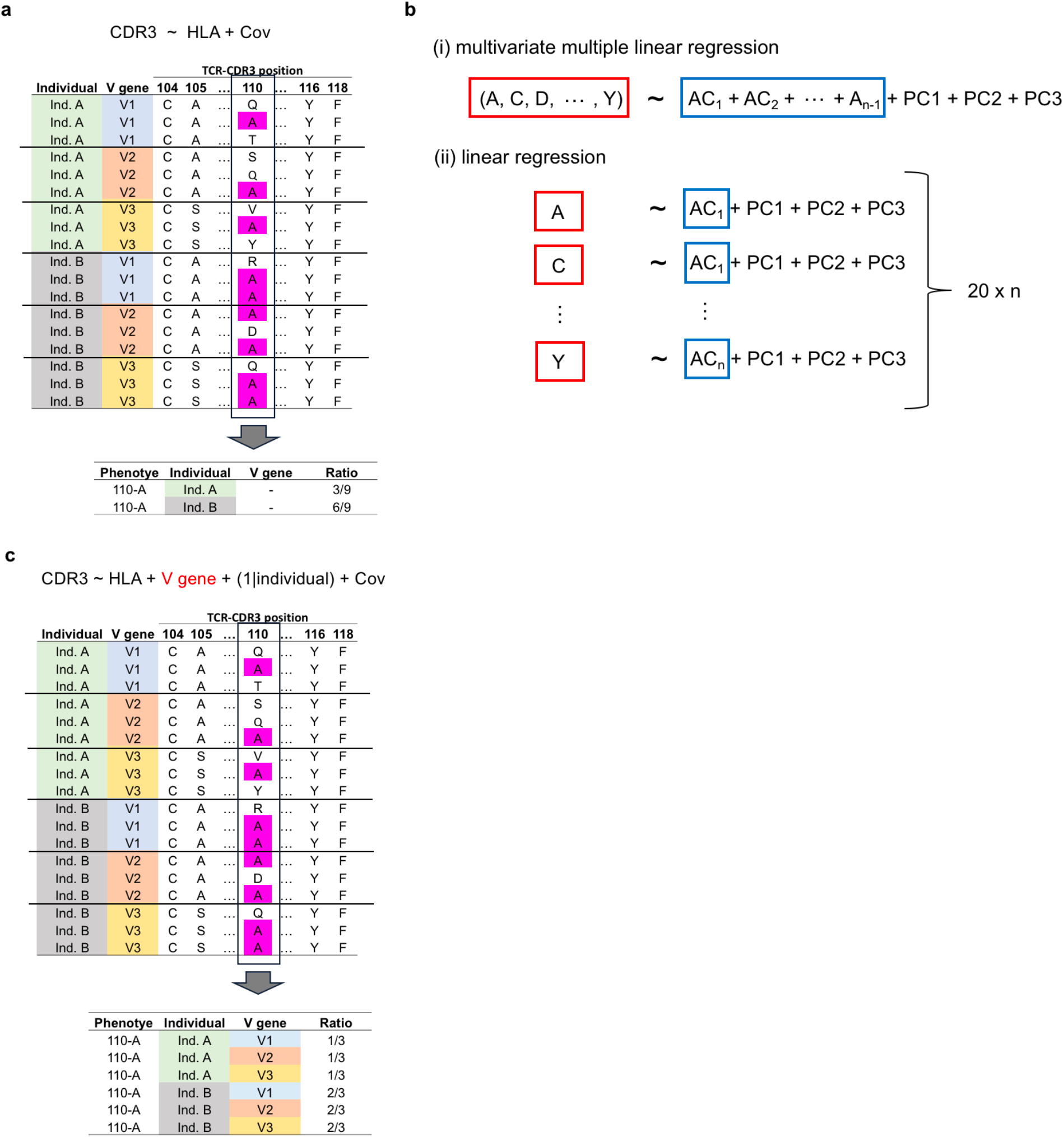
Statistical models used in this study. Schematic explanation of the linear model used in this study. **(a)** Our strategy to calculate amino acid frequencies which we utilized for the main analysis. In this example, alanine (A) usage ratio at CDR3 position 110 is calculated for each individual. **(b)** Two main linear models utilized in this study. In a multivariate multiple linear regression, a vector of frequency of 20 amino acids at a given position of CDR3 is the response variable; all amino acid alleles except one at a site of *HLA* are the explanatory variables. In a linear regression model, the frequency of a single amino acid at a position of CDR3 is the response variable; a single amino acid allele at a site of *HLA* is the explanatory variable. **(c)** Our strategy to calculate amino acid frequencies for the linear mixed model which we utilized to adjust the effect of V genes. In this example, alanine (A) usage ratio at CDR3 position 110 was calculated for each individual for each V gene. Using this frequency vector, we can assess cdr3-QTL effects independent from V genes. The same model was used for J genes.

**Extended Data Figure 3.**
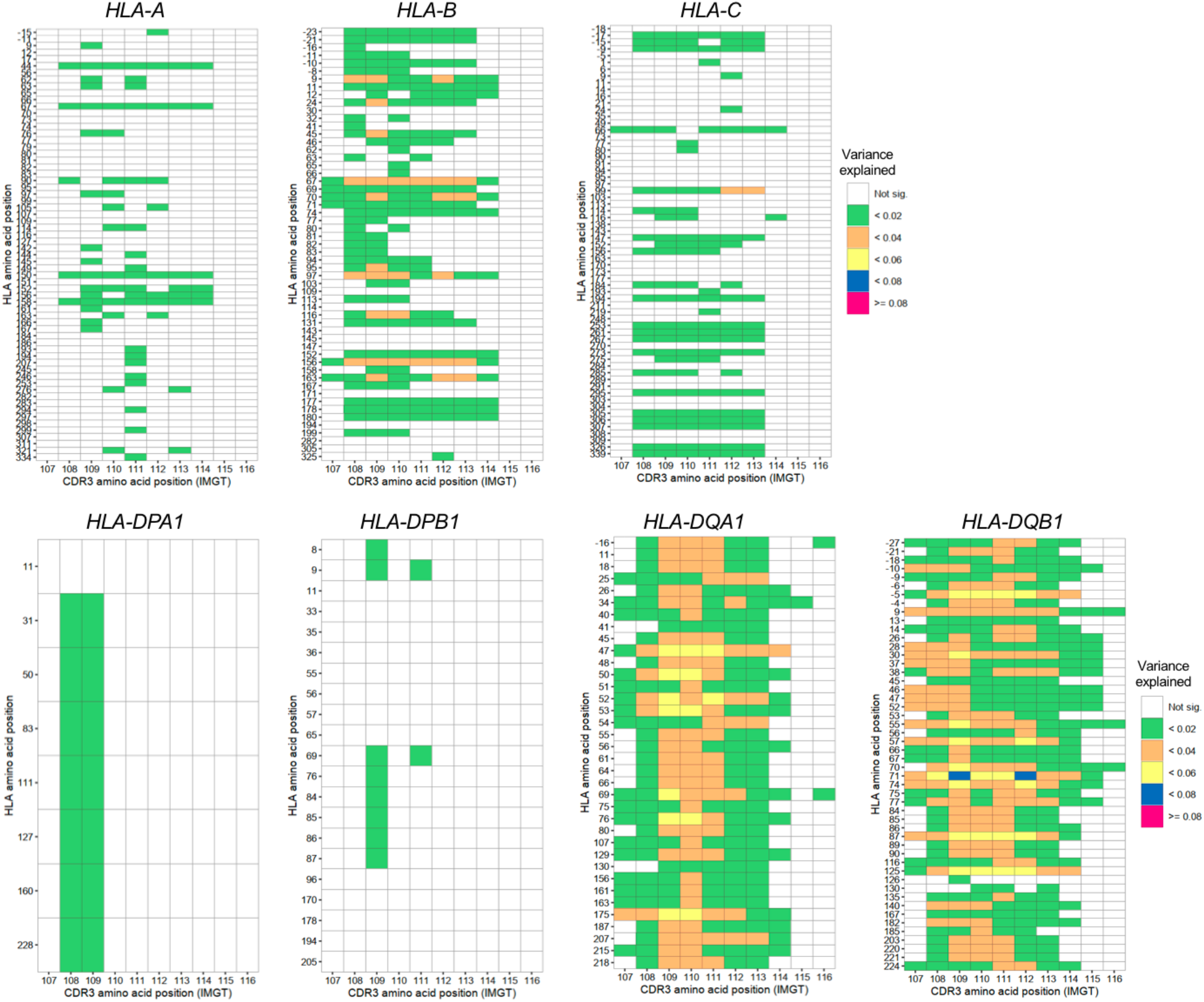
Variance explained in cdr3-QTL analysis summarized across different lengths of CDR3. Variance explained in cdr3-QTL analysis (n=628; multivariate multiple linear regression). The results for all *HLA* genes except *HLA-DRB1* were provided. For each pair of *HLA* sites and CDR3 positions, the largest variance explained across different CDR3 lengths were shown in a heatmap.

**Extended Data Figure 4.**
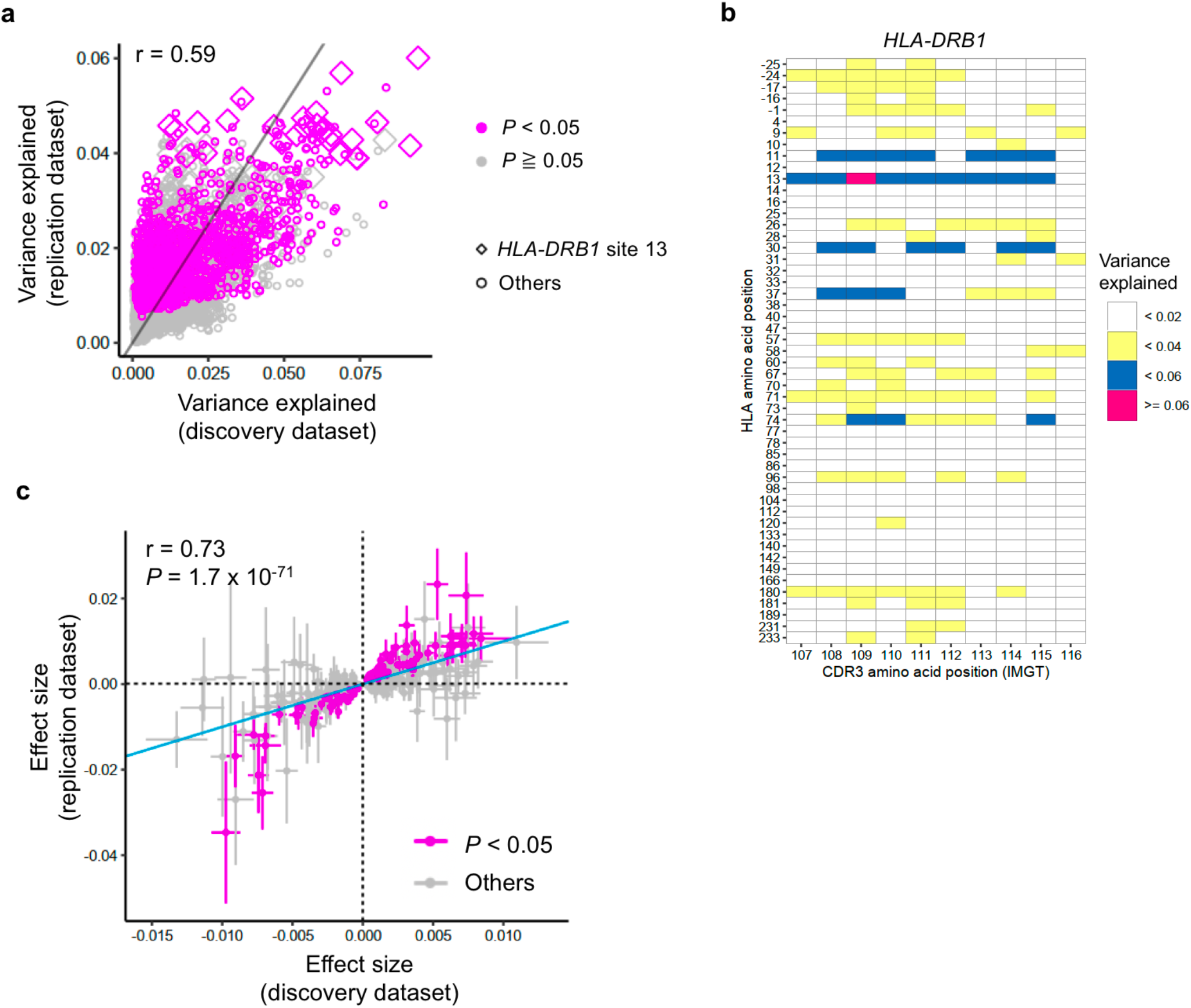
cdr3-QTL results in the replication dataset. **(a)** Explained variance in cdr3-QTL analysis in the discovery dataset (n=628; peripheral blood) was compared with those in the replication dataset (n=169; naïve CD4^+^ T cells) using multivariate multiple linear regression. All pairs of class II HLA sites and CDR3 phenotypes were shown without any filtering (n=11,550). The results at the *HLA-DRB1* site 13 and the results with *P* < 0.05 in the replication dataset were highlighted. **(b)** Explained variance in cdr3-QTL analysis in the replication dataset (multivariate multiple linear regression). For each pair of *HLA* sites and CDR3 positions, the largest variance explained across different CDR3 lengths were shown in a heatmap. The results of *HLA-DRB1* were provided. Only associations with *P* < 0.05 were colored in the heatmap. **(c)** Replication of cdr3-QTL signals using the replication dataset (n=169; a linear regression model). Effect sizes on a non-normalized scale from discovery and replication datasets were provided with S.E. The associations nominally significant in the replication dataset were highlighted by red (*P* < 0.05). The analysis was restricted to the 411 CDR3 phenotypes (length-position-amino acid combinations) which had at least one significant association in the discovery dataset (*P* < 0.05/1,262,664; linear regression) and were testable in the replication dataset.

**Extended Data Figure 5.**
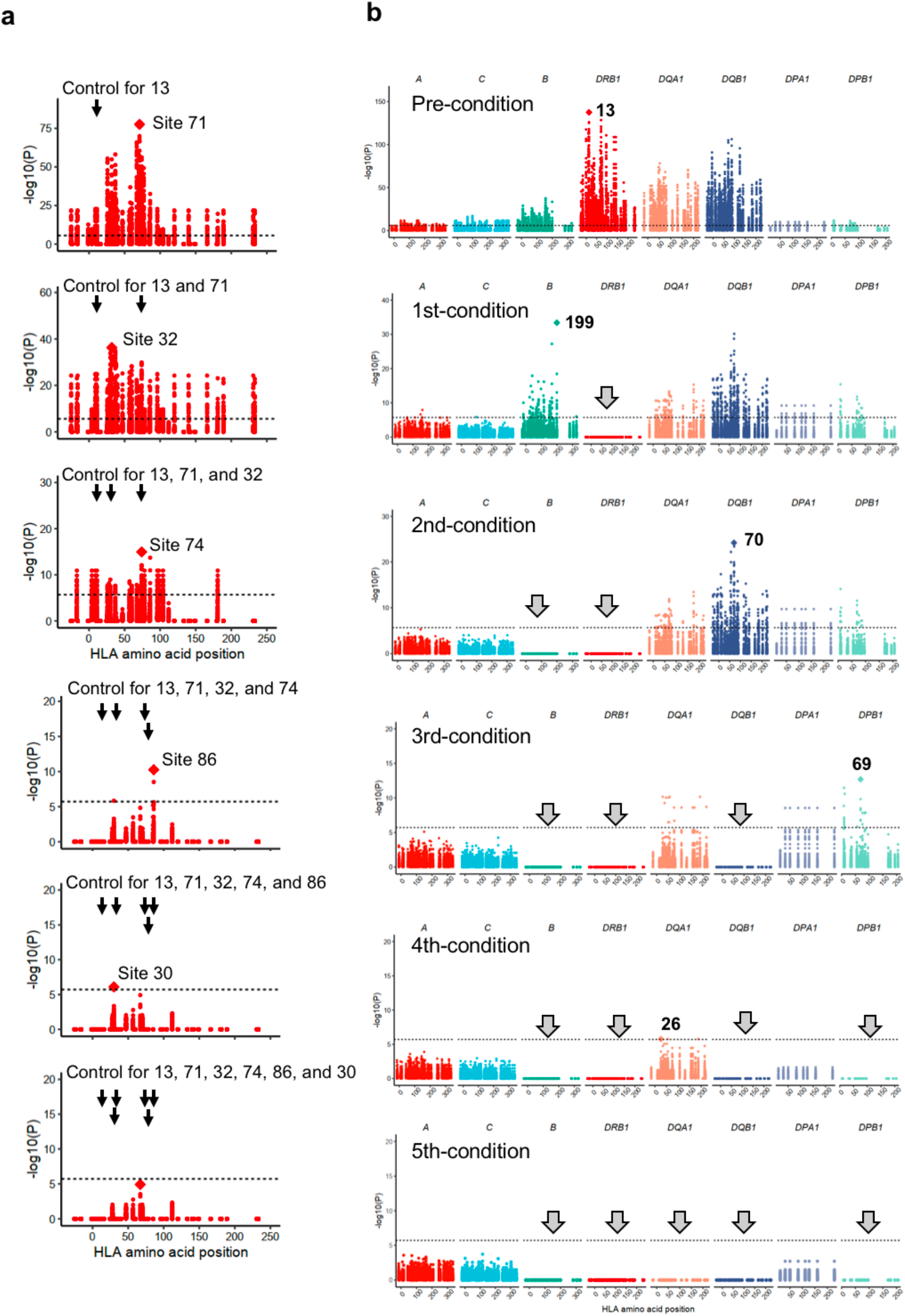
The results of conditional haplotype analysis. **(a)** Conditional haplotype analysis within *HLA-DRB1* (n=628; multivariate multiple linear regression). To test independent cdr3-QTL signals within *HLA-DRB1*, we conducted a conditional haplotype analysis using a multivariate multiple linear regression model by controlling all effects coming from specific sites of *HLA-DRB1* gene. The strongest cdr3-QTL signal was found at site 13 of *HLA-DRB1*. Therefore, in the first round of the conditional analysis, we conducted cdr3-QTL analysis by controlling the effects coming from site 13. The null model consisted of haplotypes defined only by residues at site 13. The full model consisted of haplotypes defined by the combination of residues at site 13 and the target site; addition of the target site may result in *k* additional unique haplotypes. We tested whether the creation of *k* additional haplotype groups improved the model fit. In this analysis, the strongest signal was found at site 71 of *HLA-DRB1*. Therefore, in the second round of the conditional analysis, we conducted cdr3-QTL analysis by controlling all effects coming from site 13 and 71. We repeated these processes sequentially within *HLA-DRB1* until we did not observe further significant signals (*P* > 0.05/24,430). **(b)** Conditional analysis using four-digit classical alleles (n=628; multivariate multiple linear regression). In the 1^st^ conditioning analysis, to assess whether there were independent effects outside of the *HLA-DRB1* locus, we conducted cdr3-QTL analysis using all four-digit classical alleles of *HLA-DRB1* as covariates, and the strongest signal was found in *HLA-B* region. In the 2^nd^ conditional analysis, we additionally included all four-digit classical alleles of *HLA-B* as covariates. We sequentially included all four-digit classical alleles of the gene with the strongest signal as covariates until we did not observe further significant signals (*P* > 0.05/24,430). We excluded strongly correlated alleles among covariates (r^2^ > 0.8).

**Extended Data Figure 6.**
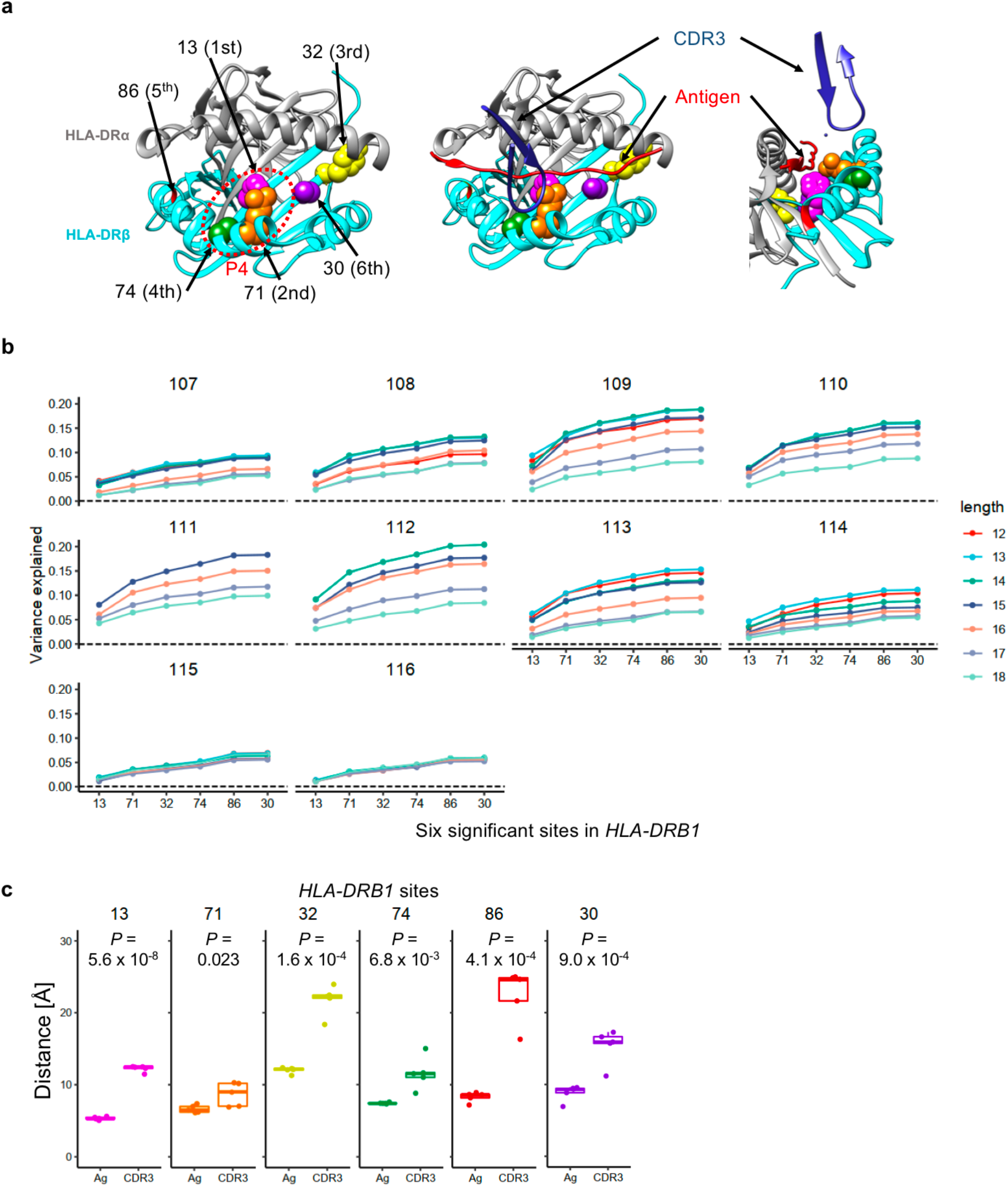
Six sites in *HLA-DRB1* with independently significant cdr3-QTL effects. **(a)** Structure of HLA-DRB1 protein and amino acid sites with independently significant cdr3-QTL effects (Protein database 2IAM). Positions 13, 71 and 74 are within the P4 binding pocket. On the left we depicted only HLA-DR molecules looking into the binding groove. In the middle, we depicted the antigen (red) and CDR3 (dark blue) overlaid onto HLA-DR molecules. On the right, we depicted HLA-DR, antigen, and CDR3 from a side view. **(b)** Variance explained by six *HLA-DRB1* amino acid sites with independently significant cdr3-QTL effects. The order of sites on the X-axis indicates the order of the significance levels. **(c)** The distances from *HLA-DRB1* amino acid sites to antigen (Ag) or to CDR3. We analyzed five structures and the shortest distances in each structure were utilized. One-sided paired t test *P* values were provided. Within each boxplot, the horizontal lines reflect the median, the top and bottom of each box reflect the interquartile range (IQR), and the whiskers reflect the maximum and minimum values within each grouping no further than 1.5 x IQR from the hinge.

**Extended Data Figure 7.**
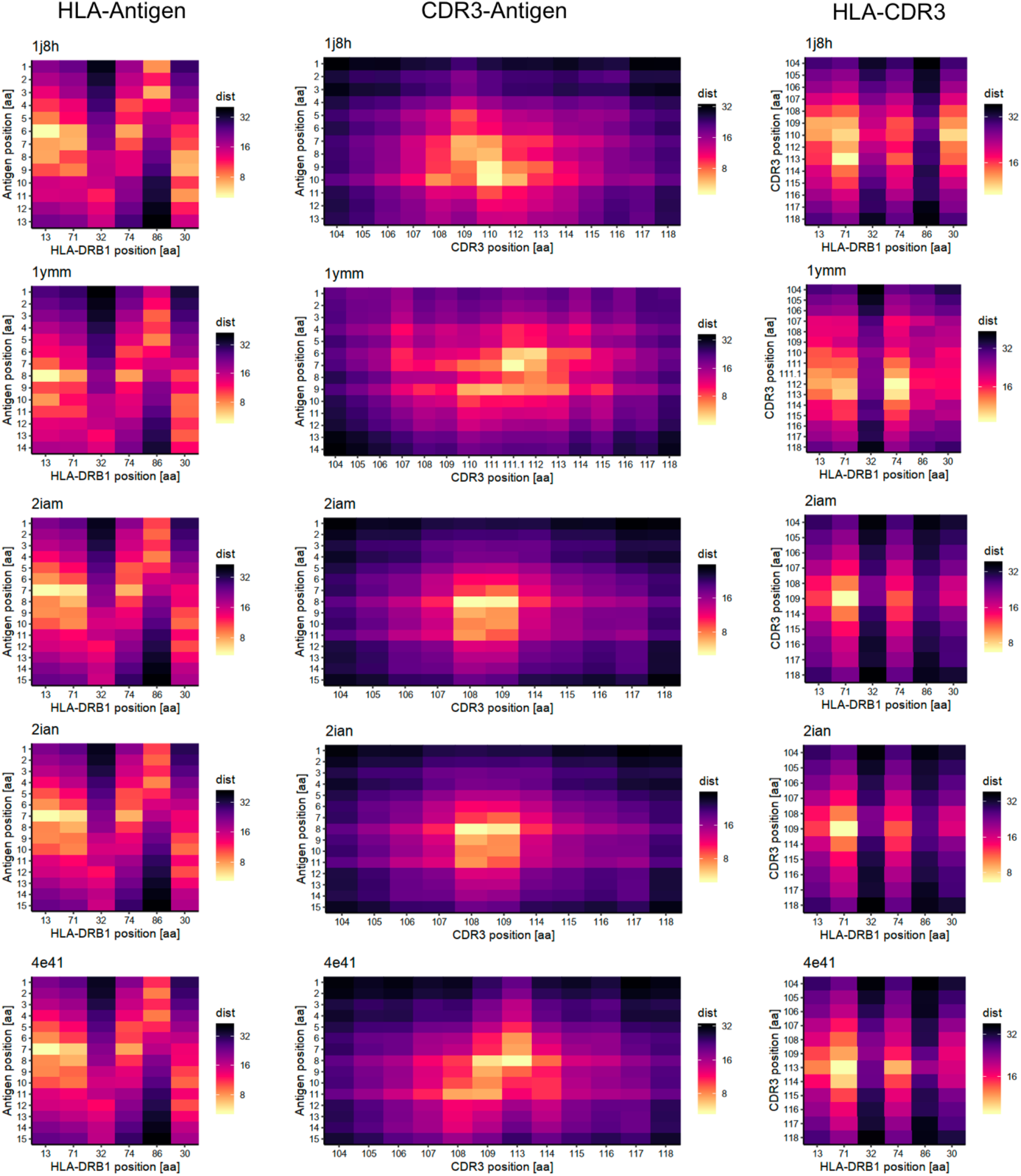
The pair-wise distances of amino acids in MHC-peptide-TCR complexes. The distances (in Å) between *HLA-DRB1* sites and antigen (left), CDR3 amino acids of beta chains and antigen (middle), and *HLA-DRB1* sites and CDR3 amino acids of beta chains (right) were shown in heatmaps.

**Extended Data Figure 8.**
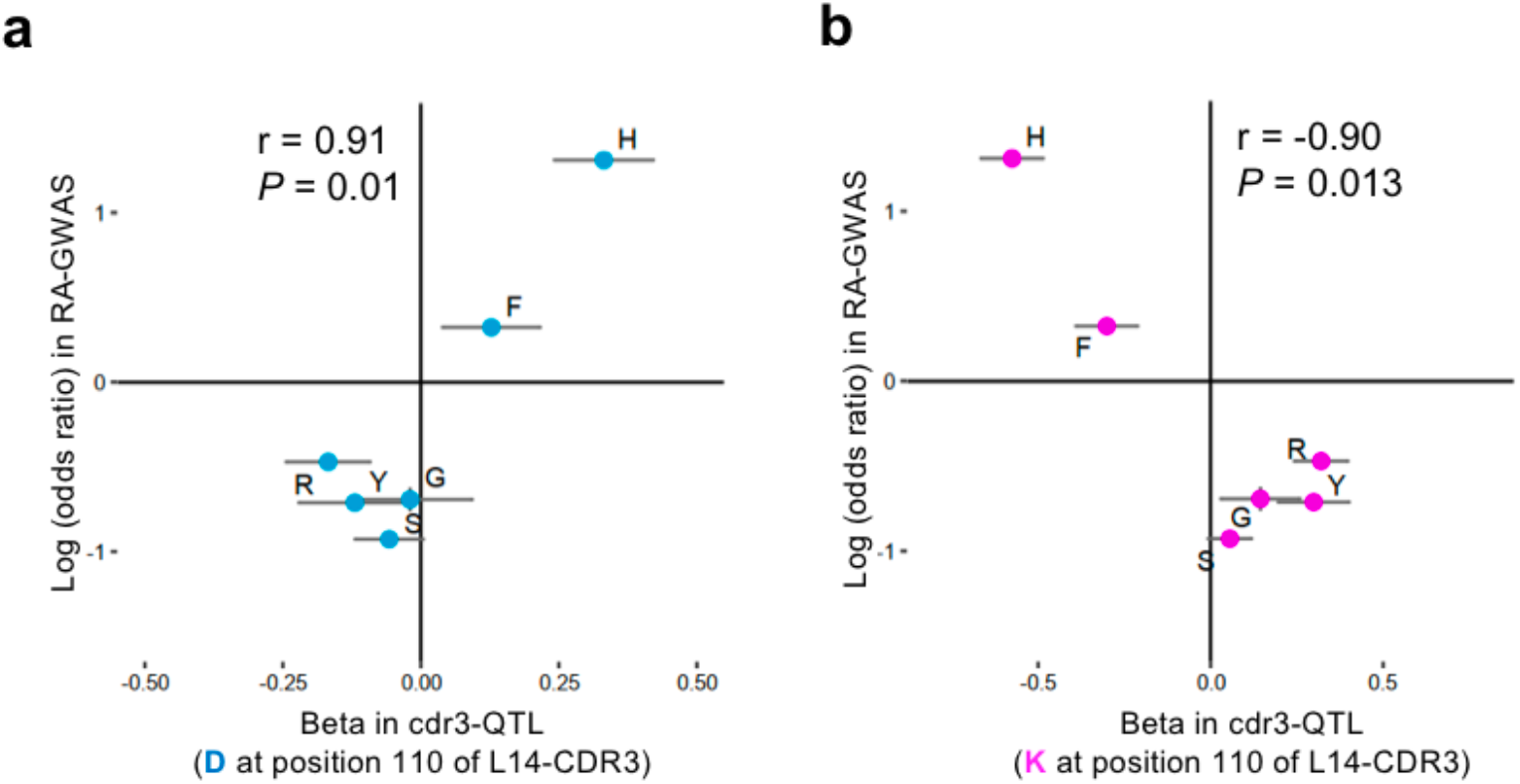
Negative charge at CDR3 position 110 might be involved in the pathogenesis of RA. **(a)** The cdr3-QTL effect sizes for aspartic acid (D) usage at position 110 of L14-CDR3 for the six possible amino acids at *HLA-DRB1* site 13 plotted against their effect size in RA-GWAS^1^ with S.E. (n=628; linear regression). **(b)** The same plot as in **a** but for lysine (K).

**Extended Data Figure 9.**
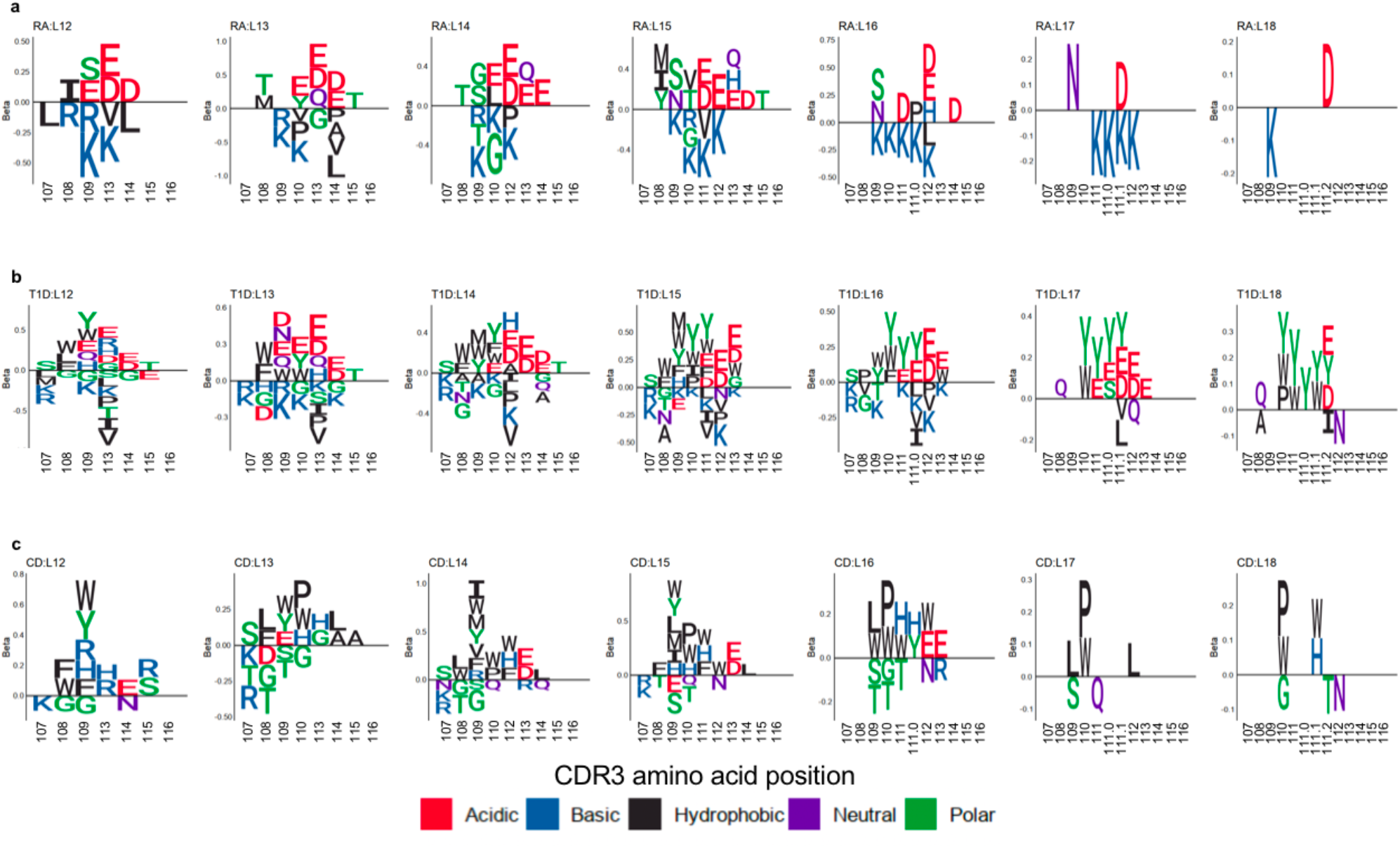
CDR3 amino acids associated with MHC-wide risk of RA, T1D, and CD. CDR3 amino acids influenced by *HLA* risk score. We conducted cdr3-QTL analysis using *HLA* risk score; the CDR3 phenotypes were each amino acid at each position of each length of CDR3 (n=628; linear regression). The effect sizes of significant associations for each amino acid at a given position were illustrated by sequence logo separately for different CDR3 lengths (**a**, RA; **b**, T1D; and **c**, CD).

**Extended Data Figure 10.**
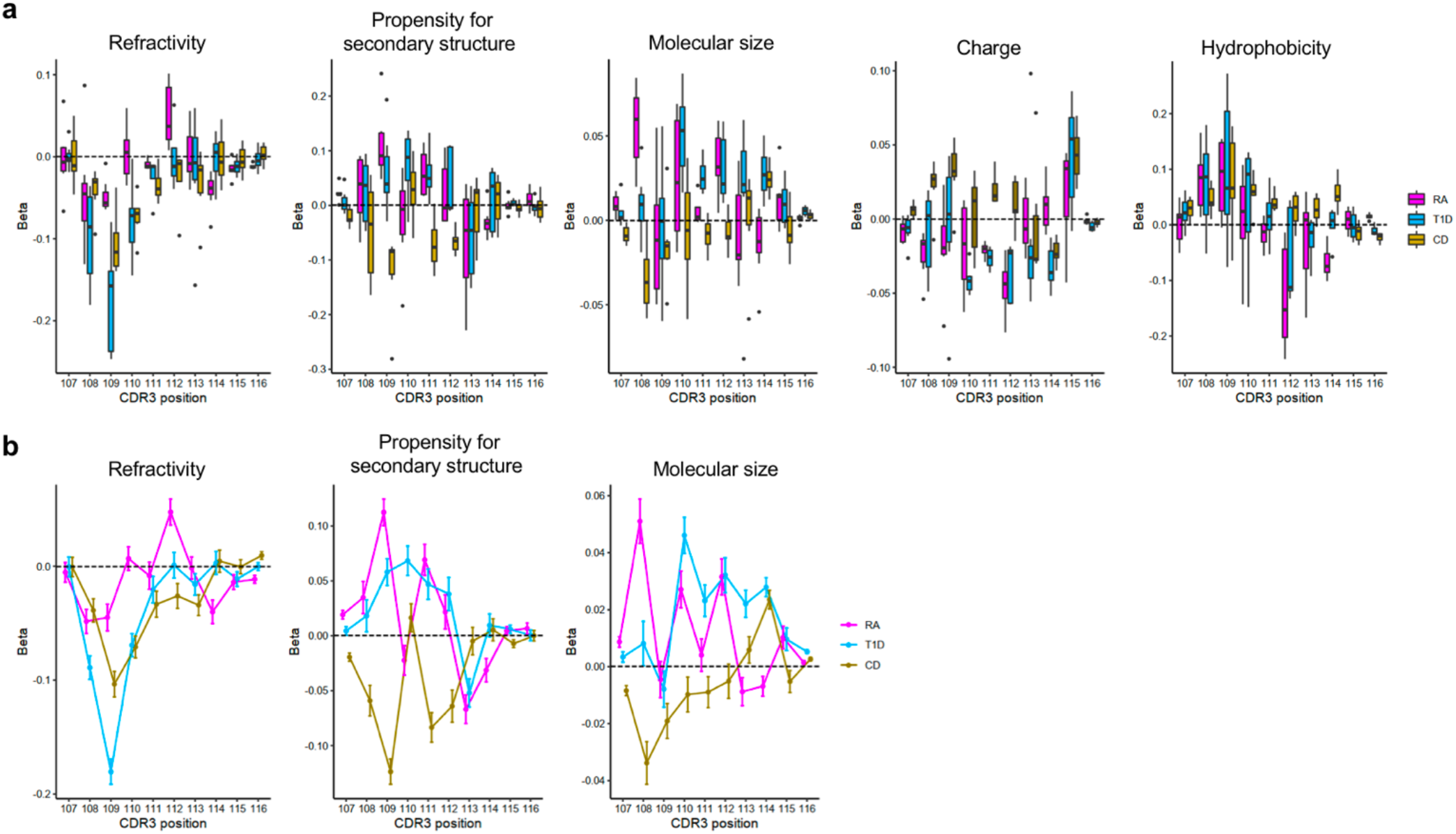
Amino acid features at each position of CDR3 influenced by *HLA* risk score. We conducted cdr3-QTL analysis using *HLA* risk score where the phenotypes were amino acid features at a given position of each length of CDR3 (n=628; linear regression). **(a)**, Effect sizes were plotted separately for different length of CDR3. **(b)**, Meta-analyzed effect sizes and S.E. were plotted (the results for charge and hydrophobicity were shown in **Figure 4c**). Within each boxplot, the horizontal lines reflect the median, the top and bottom of each box reflect the interquartile range (IQR), and the whiskers reflect the maximum and minimum values within each grouping no further than 1.5 x IQR from the hinge.

## ONLINE METHODS

### TCR-CDR3 sequencing data

We downloaded the discovery dataset from Adaptive Biotechnologies immuneACCESS site (https://clients.adaptivebiotech.com/pub/emerson-2017-natgen). The discovery dataset has CDR3 sequences of TCR beta chains from peripheral blood of healthy individuals (n = 666). This dataset has 242,461 unique CDR3 sequences per individual on average (**Table 1**). For the main analysis, we included CDR3 sequence dataset of 628 individuals whose four-digit classical alleles were available for all of *HLA-A, HLA-B, HLA-C, HLA-DQB1*, and *HLA-DRB1*. As for the demographic data, age was available for 555 samples, sex was available for 642 samples, and ethnicity data was available for 414 samples (detailed information was provided in **Table 1**). We defined amino acid sequences with stop codons or frameshifts as non-productive amino acid sequences. We utilized productive amino acid sequences of CDR3 and V/J gene information as reported in the original data. For amino acids of non-productive CDR3 sequences, we utilized MIXCR software (v2.1.11; with default parameters) since the original data did not report these sequences. For the primary analysis, we treated each unique TCR sequence as a single event irrespective of read depth to exclude the influence of clonal expansion. We considered the read depth of each TCR sequence and treated each read of TCR as a single event only when specifically discussing the effect of clonal expansion. We restricted our analysis to TCR sequences whose CDR3 has a length between 12 and 18 amino acids, which covers 94.1% of data (**Extended Data Figure 1**). We aligned CDR3 amino acids to position 104 to 118 defined by IMGT (**URLs**); CDR3 with length 12-14 have gaps in the middle and CDR3 with length 15-18 have extra positions in the middle (**Extended Data Figure 1**).

### *HLA* genotypes

We downloaded genotype data of the discovery from Adaptive Biotechnologies immuneACCESS site (https://clients.adaptivebiotech.com/pub/emerson-2017-natgen). Genome-wide genotyping data was not available for the discovery dataset, and genotype data was restricted to four-digit classical alleles of *HLA* genes. The number of samples with genotype data are different across *HLA* genes; *HLA-A* (n=629), *HLA-B* (n=630), *HLA-C* (n=629), *HLA-DPA1* (n=606), *HLA-DPB1* (n=472), *HLA-DQA1* (n=522), *HLA-DQB1* (n=630), *HLA-DRB1* (n=630). Using amino acid sequence data of classical alleles reported in IMGT (**URLs**), we identified amino acid alleles of each site of *HLA* genes. For multi-allelic amino acid positions, we defined composite markers which consist of all possible combinations of alleles in addition to each individual allele as reported in our previous studies^1,2^. Since there were many missing data for *HLA-DPA1, HLA-DPB1*, and *HLA-DQA1*, we restricted our main analysis to 628 samples whose four-digit classical alleles were available for all of *HLA-A, HLA-B, HLA-C, HLA-DQB1*, and *HLA-DRB1*, and we conducted principal component analysis (PCA) using genotypes for these five HLA genes. PCA is based on four-digit classical alleles, and we excluded correlated alleles before conducting PCA (r^2^ > 0.5).

### cdr3-QTL analysis

We treated the amino acid composition of CDR3 as a quantitative trait, and tested its association with HLA genotypes. To avoid confusion, we refer an amino acid position of *HLA* as a “site” and that of CDR3 as “position”. We analyzed each amino acid position of each length of CDR3 separately (length 12-18 amino acids). We did not analyze different positions simultaneously (i.e. we did not analyze multimers or motifs in CDR3). We excluded rare CDR3 amino acids which were observed less than 50% of individuals in a given position and length of CDR3. We also excluded rare genotypes that have a minor allele frequency (MAF) less than 1%. We conducted cdr3-QTL analysis using three different models as follows (**Extended Data Figure 2**).

#### i) multivariate multiple linear regression

This model was utilized to quantify the correlation between multiple amino acids at a position of CDR3 and multiple amino acid alleles at a site of HLA. We calculated the proportion of the unique CDR3 sequences that had each of the twenty amino acid residues at a given position of CDR3, and we used this information to create a twenty-dimensional phenotype vector. For each component of this phenotype vector, proportions were transformed into a standard normal distribution across individuals (rank-based). At a given HLA site which has *m* amino acid residues, we partitioned the classical alleles into *m* groups with identical residues at that site, and calculated the allele dosage of each group. We included top three principal components of genotypes in this analysis. As a result, the full model is the following multivariate multiple linear regression model:

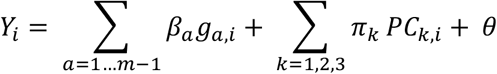

where *Y*_*i*_ indicates twenty-dimensional phenotype vector of individual *i, a* indicates a specific group of classical alleles being tested, and *g*_*a,i*_ is the dosage of allele group *a* in individual *i*. The *β*_*a*_ is a twenty-dimensional parameter which represents the additive effect per allele. We included *m* − 1 groups of classical alleles, excluding 1 group as a reference. The *π*_*k*_ is a twenty-dimensional parameter which represents the effect for the *k*-th principal components, and *PC*_*k,i*_ is the value for individual *i* for the *k*-th principal component. The θ is a twenty-dimensional parameter which represents the intercept.

The null model only had terms for covariates without allelic effects:

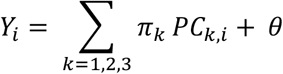

We estimated the improvement in the model fit between the full model and the null model using MANOVA, and assessed the significance of the improvement using Pillai statistics. The variance of twenty-dimensional phenotype vector explained by *m* − 1 genotype groups was estimated using R package MVLM. We calculated the variance explained by full and null models separately, and we defined the variance explained by genotypes as the difference between the two values.

#### ii) linear regression

This model was utilized to quantify the correlation between each amino acid at a position of CDR3 and each amino acid allele at a site of *HLA* gene. We utilized the same phenotypes (but one-dimensional phenotype in this model) as utilized in the multivariate multiple linear regression model. We utilized amino acid allele dosages at each *HLA* site as explained above.

We utilized the following linear regression model:

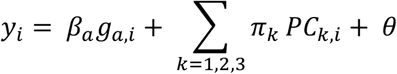

where *y*_*i*_ indicates a scaler phenotype of individual *i, a* indicates a specific *HLA* allele being tested, and *g*_*a,i*_ is the dosage of allele *a* in individual *i*. The *β*_*a*_ is a scaler parameter which represents the additive effect per allele *a*. The *π*_*k*_ is a scaler parameter which represents the effect for *k*-th principal components, and *PC*_*k,i*_ is the value for individual *I* for the *k*-th principal component. The θ is a scaler parameter which represents the intercept.

#### iii) linear mixed regression

This model is similar to the linear regression model, but includes an additional fixed effect term for V genes and a random effect term for individuals:

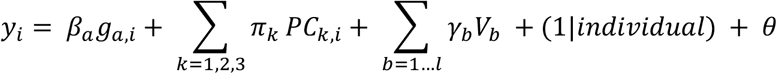

where the *γ*_*b*_ is a parameter which represents the effect for V gene *b*, and *V*_*b*_ is an indicator variable that is equal to 1 only if the CDR3 has V gene *b*. Since adding a V gene term creates multiple observations from each individual, we included a random effect term for the individual to control for the repeated measurements. The definition of other terms is the same as those in the linear regression model. We utilized R package lme4.

### *HLA* gene-level conditional analysis

To test independent cdr3-QTL signals outside of a given *HLA* gene, we conducted a conditional analysis using a multivariate multiple linear regression model by controlling all effects coming from that *HLA* gene. The strongest cdr3-QTL signal was found within the *HLA-DRB1* locus. Therefore, in the first round of the conditional analysis, we conducted cdr3-QTL analysis using all four-digit classical *HLA-DRB1* alleles as covariates. In this analysis, the strongest signal was found within the *HLA-B* locus. Therefore, in the second round of the conditional analysis, we conducted cdr3-QTL analysis using all four-digit classical alleles of *HLA-DRB1* and *HLA-B* as covariates. We repeated these processes sequentially until we did not observe further significant signals (*P* > 0.05/24,430). We excluded strongly correlated alleles among covariates (r^2^ > 0.8).

### *HLA* site-level conditional analysis

To test independent cdr3-QTL signals within a given *HLA* gene, we conducted a conditional haplotype analysis using a multivariate multiple linear regression model by controlling all effects coming from specific sites of that *HLA* gene. The strongest cdr3-QTL signal was found at site 13 of *HLA-DRB1*. Therefore, in the first round of the conditional analysis, we conducted cdr3-QTL analysis by controlling the effects coming from site 13. The null model consisted of haplotypes defined only by residues at site 13. The full model consisted of haplotypes defined by the combination of residues at site 13 and the target site; addition of the target site may result in *k* additional unique haplotypes if the site is independent from site 13. We tested whether the creation of *k* additional haplotype groups improved the model fit. In this analysis, the strongest signal was found within site 71 of *HLA-DRB1*. Therefore, in the second round of the conditional analysis, we conducted cdr3-QTL analysis by controlling the effects coming from site 13 and 71. We repeated these processes sequentially within *HLA-DRB1* until we did not observe further significant signals (*P* > 0.05/24,430).

### Structural analysis of MHC-peptide-TCR complex

We downloaded the structural analysis results of MHC-peptide-TCR complexes from Protein Data Bank (**URLs**). We restricted our analysis to the results which analyzed *HLA-DRB1* and had all positional data for all three molecules: the HLA proteins, the antigenic peptide, and the TCR beta chain. These are Protein Data Bank entries 1J8H, 1YMM, 2IAM, 2IAN, and 4E41. For each amino acid, we first calculated the centroids of every atom using the XYZ orthogonal Å coordinates reported in the database; and we next calculated the pair-wise distances between amino acids using the centroid positions.

We calculated pairwise distances between HLA, TCR, and antigen amino acids and embedded them into a two-dimensional space. In this embedding, we preserved distances important for antigen recognition; distances between HLA-DRB1 and antigens and those between TCR and antigens, and down-weight the distances between HLA-DRB1 and TCR (**Supplementary Note**).

We visualized the *HLA-DRB1* amino acid sites which had independently significant associations in conditional haplotype analyses. We utilized UCSF Chimera software to create a 3-D view of the MHC-peptide-TCR complex based on Protein Data Bank entries 2IAM.

### Replication analysis using naïve CD4^+^ T cells

All replication dataset was generated by BLUEPRINT consortium and was downloaded from European Genome-phenome Archive under the accession code of EGAD00001002671 and EGAD00001002663 (**URLs**). We analyzed FASTQ files of RNA sequencing data of naïve CD4^+^ T cells (n = 169 healthy donors). Demographic data was provided in **Table 1**. Reads were mapped to GRCh38 human reference sequences with GENCODE v26 gene models by STAR software (v2.5.3). Using reads mapped to the *TCR* loci (chr7: 142,299,011-142,813,287 for beta chain) and unmapped reads, we analyzed TCR sequences using MIXCR (v2.1.11; with default parameters). Using genotype data around the *MHC* locus, we imputed *HLA* genotypes by SNP2HLA software using T1DGC reference panel (n= 5225 European samples), and excluded poor quality genotypes (r^2^ < 0.5). We conducted PCA by Plink software (v1.90) using LD-pruned genome-wide variants (r^2^ > 0.2).

### *HLA* risk score

To evaluate MHC-wide risk of RA, T1D, and CD for each individual, we defined an *HLA* risk score for each disease. First, we defined critical haplotypes of *HLA* amino acids which confer disease risk based on the results of previous studies^1,2,29^ (**Supplementary Table 9**). For RA, we included four amino acid sites; *HLA-DRB1* sites 11, 13, 71, and 74. For T1D, we included three amino acid sites; *HLA-DQB1* site 57, and *HLA-DRB1* sites 13 and 71. For CD, we included four-digit classical alleles of *HLA-DRB1* and *HLA-DQB1*. Since many samples did not have *HLA-DQA1* genotypes (n=144), we did not include the genotype of *HLA-DQA1*. We also calculated the odds ratio of each haplotype according to multivariate regression in previous GWAS (**Supplementary Table 9**). We then calculated the product of effect sizes (= log(odds ratio)) and dosages of those haplotypes for each individual, and the sum of two products was defined as *HLA* risk score of an individual. The *HLA* risk score has a different distribution for each disease. When we conducted regression using *HLA* risk score, we scaled the HLA risk scores so that its S.D. is equal to 1, and hence effect sizes of *HLA* risk score of different diseases are comparable with each other.

### Amino acid features

Amino acids have multiple complex physiochemical and biological properties. To analyze amino acid features comprehensively, we utilized previously-reported five numeric features which summarize the entire constellation of amino acid physiochemical properties (each amino acid has a unique value for each feature). Briefly, these five features were derived from factor analysis based on 494 amino acid indices, which include general attributes (e.g. molecular volume) as well as more specific measures (e.g. side chain orientation angle). Based on the original report, we annotated factor I as charge, factor II as hydrophobicity, factor III as refractivity, factor IV as propensity for secondary structure, and factor V as molecular size. For factor II, the original value indicated hydrophilicity, and we flipped the sign so that the value represents hydrophobicity. When we conducted cdr3-QTL analysis using these amino acid features, we calculated weighted average of a given amino acid feature as a phenotype; we multiplied the frequency of each amino acid by its value for a given feature, and calculated the sum of the product. When we conducted cdr3-QTL analysis for a given feature, we added the other four features as covariates to handle correlations between these features.

### CDR3 risk score

We developed a scoring system which indicates the extent of each CDR3 sequence’s association with *HLA* risk score, and we refer to this as CDR3 risk score. This is analogous to polygenic risk score (PRS). We utilized effect sizes of *HLA* risk score on each amino acid residue in each position of CDR3 with each length (L12-18) using a linear regression model (i.e. effect sizes of cdr3-QTL analysis based on *HLA* risk score). The risk score is the sum of effect sizes of amino acids in a given CDR3 sequence, specified to both the position of each amino acid and the overall CDR3 length (**Supplementary figure 8**).

As in PRS, *P* value threshold in cdr3-QTL analysis is an important tuning parameter for CDR3 risk score. Therefore, we determined an appropriate *P* value threshold with five-fold cross validation in the discovery dataset. In each round of cross validation, we conducted cdr3-QTL analysis based on *HLA* risk score using 80% of samples, and we utilized the effect sizes from this analysis to calculate CDR3 risk score in the testing samples (the remaining 20% of samples). The CDR3 risk score is expected to be correlated with *HLA* risk score based on its definition. Therefore, we evaluated the performance of CDR3 risk score by using its correlation with *HLA* risk score. Using RA-CDR3 risk score, we confirmed that Bonferroni-corrected *P* value (= 0.05/1,368) had the best performance (**Supplementary Figure 8**). Therefore, for the main analysis of the CDR3 risk score, we included the 94, 206, and 123 CDR3 phenotypes (length-position-amino acid combinations) for RA, T1D, and CD, respectively that passed Bonferroni-corrected *P* value threshold (P < 0.05/1,368).

### TCR sequences of tetramer-positive T cells

We analyzed sequence data of TCRs that were reactive to candidate pathogenic epitopes for RA and CD. We restricted our analysis to CDR3s with a length between 12 and 18 amino acids, and calculated CDR3 risk scores for RA and CD. We utilized one-sided t test to assess the significance of difference in CDR3 risk scores between two groups.

For the analysis of RA, we collected peripheral blood of RA patients (n=7 in total) and expanded T cells using peptides corresponding to relevant citrullinated epitopes^41,42^. We conducted fluorescence-activated cell sorting (FACS) using HLA-DRB1*0401- or HLA-DRB1*0404-tetramer loaded with citrullinated aggrecan (cit-aggrecan) or citrullinated CILP (cit-CILP), isolating and expanding single cells and then sequencing the TCRs of these T cells^43^. We thus identified six cit-aggrecan-specific TCR sequences from five RA patients and five cit-CILP-specific TCR sequences from two RA patients.

Patients in this analysis had at least one allele of *HLA-DRB1*\*0401 or *HLA-DRB1*\*0404, though their *HLA* genotypes were not necessarily identical. Since we did not have control TCR sequences from the same patients, we utilized TCR sequences from the replication dataset (naïve CD4T cells in peripheral blood of healthy individuals). We restricted the samples in the replication dataset to individuals who were homozygous for *HLA-DRB1*\*0401 (the allele with the highest *HLA* risk score). Since CDR3 risk score is positively correlated with HLA risk score by definition, this strategy should identify individuals who have TCR repertoire with the highest CDR3 risk score; and hence this strategy is a conservative approach. One individual who has 1,753 TCR sequences met this criterion.

For the analysis of CD, we searched the literature for studies that utilized tetramers to identify gliadin epitope-specific CD4^+^ T cells and reported their sequences. Three studies met these criteria^36–38^, and included seven TCRs specific to α-I gliadin (n = 4 patients), 92 TCRs specific to α-II gliadin (n = 13 patients), eight TCRs specific to ω-II gliadin (n = 2 patients), and 49 control TCRs (n = 3 patients). Patients in these reports had at least one HLA-DQ2 haplotype (*HLA-DQA1*\*05:01-*HLA-DQB1*\*02:01), though their *HLA* genotypes were not necessarily identical.

### Data availability

All raw TCR sequence data and genotype data of the discovery dataset and the replication dataset are available at Adaptive Biotechnologies immuneACCESS site (https://clients.adaptivebiotech.com/pub/emerson-2017-natgen) and European Genome-phenome Archive under the accession code of EGAD00001002671 and EGAD00001002663 (URL). All summary statistics of cdr3-QTL analysis are available at our website (https://github.com/immunogenomics/cdr3-QTL).

### Code availability

All code utilized in this study is available at our website (https://github.com/immunogenomics/cdr3-QTL).

## Data Availability

All raw TCR sequence data and genotype data of the discovery dataset and the replication dataset are available at Adaptive Biotechnologies immuneACCESS site and European Genome-phenome Archive under the accession code of EGAD00001002671 and EGAD00001002663. All summary statistics of cdr3-QTL analysis are available at our website.

## URL

Adaptive Biotechnologies immuneACCESS site: https://clients.adaptivebiotech.com/

European Genome-phenome Archive: https://www.ebi.ac.uk/ega/home

THE INTERNATIONAL IMMUNOGENETICS INFORMATION SYSTEM (IMGT): http://imgt.org/

Protein database: https://www.rcsb.org

GWAS Catalog: https://www.ebi.ac.uk/gwas/

## ACKNOWLEDGEMENT

This work was supported in part by funding from the National Institutes of Health (AR063759-05 (SR), U01-HG009379-04 (SR), U19-AI111224-06 (SR), T32GM007753 (KL)). K.I was supported by The Uehara Memorial Foundation. We also acknowledge Drs. Michael Brenner, A. Helena Jonsson, and Deepak Rao for helpful feedback.

## AUTHOR CONTRIBUTIONS

K.I. and S.R. conceived and designed the study. K.I. conducted all analyses with support from K.L, Y.L, and S.R. E.J. and J.B generated and managed TCR data from RA patients. K.I. and S.R. wrote the initial draft of the manuscript. All co-authors contributed to the final manuscript.

