## Supplementary note and figures for "*HLA* autoimmune risk alleles restrict the hypervariable region of T cell receptors"

### Replication analysis using a multivariate multiple linear regression model

To reproduce the results from the discovery dataset, we obtained a replication data set of 169 healthy individuals consisting of RNA-seq data from sorted naïve $CD4^+$ T cells (**Table1**)[1]. This dataset offered some advantages; the presence of only $CD4^+$ T cells; the use of naïve T cells excluded antigen experienced memory T cells; and all individuals were from the same continental ancestry (European populations) and genotyped genome-wide. Therefore, in this analysis we were able to strictly control for the effects of population stratification using principal components of genome-wide genotype data. For this dataset, we inferred TCRs from bulk RNA-seq, and hence there were fewer CDR3 sequences per individual than in the discovery dataset (around 0.7% of those observed in the discovery dataset; **Table 1**), resulting in missing low-frequency CDR3 amino acid residues in certain instances and reduced power. We applied the same analysis to the replication dataset. From 24,430 tests in the discovery dataset, we selected 11,620 tests within *HLA* class II since we only had $CD4^+$ T cells in the replication dataset; among those the vast majority (n=11,550) were frequent enough to test for association (see **Methods**). We observed that the variance explained in each test was similar in the replication dataset as in the discovery dataset (Pearson's r = 0.59), and the largest explained variance was again identified at *HLA-DRB1* site 13 and L13-CDR3 position 109 (**Extended Data Figure 4**).

### Replication analysis using a linear regression model

In our replication data, we sought to test the replicability of the 435 significant CDR3 phenotypes (length-position-amino acid combinations) at *HLA* amino acid alleles which showed the strongest association in the discovery dataset. Out of these 435 phenotypes, a total of 421 phenotypes were testable in the replication dataset (some HLA alleles and CDR3 phenotypes were missing due to low frequencies in the replication dataset). Among them, we only tested the 411 phenotypes whose lead associations located in the class II HLA since we only had $CD4^+$ T cells in the replication dataset. The effect sizes from the discovery and replication dataset were significantly correlated (r = 0.73; $P = 1.7 \times 10^{-71}$; **Extended Data Figure 4; Supplementary Table 8**); 344 of 411 phenotypes were replicated in the same allelic direction (sign test $P$ value = $5.7 \times 10^{-46}$). If we restrict this analysis to 92 phenotypes for which we found nominally significant associations in the replication dataset ($P < 0.05$), all of them replicated in the same allelic direction (sign test $P$ value = $4.0 \times 10^{-28}$). Therefore, the majority of replication failures might be due to insufficient statistical power of the replication dataset which has a smaller sample size and much less CDR3 sequences than the discovery dataset.

**Thymic selection may be driving HLA-CDR3 associations**

Since we observed consistent cdr3-QTL signals in PBMCs (including naïve and memory T cells) and sorted naïve T cells, we hypothesized that cdr3-QTL effects might be driven by thymic selection. Alternative possibilities included that cdr3-QTLs were driven by genetic mechanisms prior to thymic selection, or by antigen presentation in the periphery by *HLA* alleles.

To investigate the possibility of a genetic mechanism prior to thymic selection, we analyzed non-productive CDR3 sequences. Although they are generated by the same random recombination as productive CDR3s, they are not expressed on T cell surfaces and thus are not subjected to thymic selection. We predicted that if thymic selection is driving cdr3-QTLs, we should not observe HLA-CDR3 associations in non-productive sequences. Indeed, when we tested individual *HLA* sites to assess if they explained the variance of CDR3 amino acid frequencies at each position (multivariate multiple linear regression), we observed no significant signals in non-productive sequences (minimum $P = 3.7 \times 10^{-5} > 0.05/24{,}430$; **Figure 3a**). Since non-productive sequences were only 17.9% of all unique CDR3 sequences, we considered the possibility that this reflects reduced power. However, when we downsampled the productive sequences to match the number of non-productive sequences and repeated the analysis, we observed that productive CDR3s had substantial evidence of cdr3-QTL (**Figure 3a**). Consistently, when we analyzed individual amino acids in non-productive sequences using a linear regression model, there was no consistency in effect size directions with productive sequences, suggesting non-productive CDR3 sequences produced random results (**Figure 3b**).

If peripheral antigen presentation by MHC and memory formation was ultimately driving the observed effects, then T cells with CDR3 favored by specific *HLA* alleles would be expanded due to proliferation. In this case, weighting each unique CDR3 sequence by its expansion level should augment cdr3-QTL signals, relative to our primary analysis in which we treated each unique sequence equally without any weights. To test this possibility, we reanalyzed the discovery data; we included the effect of clonal expansion by considering the read counts of each CDR3 sequence. We still observed evidence of cdr3-QTL effects but with a substantially lower magnitude (**Figure 3c-d**). In addition to the replication analysis results using naïve T cells, these results suggested that our cdr3-QTL results reflect the effects of thymic selection favoring individual CDR3 sequences, and that these signals are mitigated (rather than augmented) by peripheral clonal expansion.

#### Identifying cdr3-QTL loci outside of the *MHC* regions

Since there is some correlation between V/J genes and CDR3 amino acids, we considered that cis-regulatory variants of V/J genes within the *TCR* locus might affect CDR3 amino acid composition indirectly. In the replication dataset for which genome-wide genotype data was available, we searched for the cis-regulatory variants of V/J genes and CDR3 amino acid compositions of beta chains among the 940 variants in the *TCR* locus (Chr. 7:141,998,851-142,510,972 in the GRCh37 genomic coordinates). Among 48 V and 13 J genes we detected, we observed significant associations for 22 V genes and nine J genes ($P < 8.7 \times 10^{-7} = 0.05 / (940 \times (48 + 13))$; **Supplemental Table 2** and **Supplementary Figure 1**). In addition, among 1,020 CDR3 phenotypes (length-position-amino acid combinations; potentially 7 x 10 x 20 = 1,400 phenotypes but we could not detect rare amino acid in the replication dataset), we observed significant associations for 47 phenotypes ($P < 5.2 \times 10^{-8} = 0.05 / (940 \times 1{,}020)$; **Supplementary Table 3** and **Supplementary Figure 1**). Of the 1,547 significant associations, 1,461 (94.4%) were to positions 113-116 just adjacent to J genes. Accounting for nine J genes with significant cis-regulatory variants completely obviated associations with CDR3 amino acid composition (**Supplementary Figure 1**). These results suggested that cis-regulatory effects for CDR3 amino acid compositions are mainly driven by their correlation with J genes and limited to the flanking positions of CDR3. Therefore, effects of cis-regulatory variants are likely distinct from those of HLA alleles, which mainly influence the middle positions of CDR3 located closer to antigens.

#### CDR3 risk score

CDR3 risk score is analogous to genetic risk score; the effect sizes of cdr3-QTL analysis based on *HLA* risk score (this is the identical analysis as we summarized in **Figure 4b**) were summed up when the target amino acid exists in a given CDR3 sequence (**Supplementary Figure 8** and **Methods**). For the CDR3 risk score, the choice of *P* value threshold can be flexible, and the one with the best performance should be utilized for the downstream analysis; we defined the performance of CDR3 risk score by the correlation between the average CDR3 risk scores and the *HLA* risk scores. Using 5-fold cross validation in the discovery dataset, we tested five different *P* value thresholds; 0.05, 0.01, 0.001, $1 \times 10^{-4}$, $3.6 \times 10^{-5}$ (= 0.05/1,368, Bonferroni corrected *P* value). We decided to choose a *P* value threshold of $3.6 \times 10^{-5}$ since we confirmed that the score with this threshold had the best performance in the cross validation (**Supplementary Figure 8**).

**Embedding of TCR 3-D structure**

In order to embed points into a two-dimensional space, we obtained protein structure data on TCR, HLA-DR, and antigen structures (1J8H, 1YMM, 2IAM, 2IAN, and 4E41). Using the centroids of each amino acid residue, we calculated distances, $d_{i,j}$, between amino acid residues *i* and *j*, from HLA-DRB1 and antigen, antigen and CDR3, and HLA-DRB1 and CDR3. We examined only polymorphic HLA residues. We averaged distances across all five structures.

If a residue centroid had an averaged pairwise distances >20 angstroms for all measured distances, we did not include those residues in our analyses. For remaining points, we sought to embed the centroids into a two-dimensional plot, by assigning x and y coordinates in order to minimize the difference between distances in embedded space, and distance in three-dimensional structural space. For each pairwise distance, we used a weight $w_{ij}$ to emphasize certain distances, and de-emphasize others. We sought to minimize the following objective function:

$$F(\vec{x},\vec{y}) = \sum_{i=1}^{n-1}\sum_{j=i+1}^{n} w_{i,j}\left(\sqrt{\left(x_i - x_j\right)^2 + \left(y_i - y_j\right)^2} - d_{i,j}\right)^2$$

To insure short distances (d<6 angstroms), and de-emphasize longer distances, we defined $w_{ij}$ as follows:

$$w_{i,j} = f(x) = \begin{cases} 5, & if\ d_{ij} < 6, one\ of\ i,j\ is\ from\ antigen \\ \max\left(0.1, (4.1667) * \left(1 - \frac{d_{ij}}{15}\right)\right), & if\ 6 \le d_{ij} < 20, one\ of\ i,j\ is\ from\ antigen \\ 2.5, & if\ d_{ij} < 6, i,j\ are\ from\ TCR\ and\ HLA - DR \\ \max\left(0.05, (2.08335) * \left(1 - \frac{d_{ij}}{15}\right)\right), & if\ 6 \le d_{ij} < 20, i,j\ are\ from\ TCR\ and\ HLA - DR \end{cases}$$

To keep residues from collapsing in on themselves, we make certain arbitrary assignments. If *i,j* are from two residues that are from within the same molecule, or are from two residues molecules that are >20 angstroms, we assign the $d_{i,j}$ = 50 angstroms, and w(i,j) to be 0.01.

To identify the best fit, we used random start positions, where each point was randomly assigned to twenty points. After a random starting point, we optimized the fit of the points by using both gradient descent and Newton's method. Newton's method needs more computational time per iteration, since the Jacobian of F needed to be calculated. Therefore, we applied 90 iterations of gradient descent first:

$$\begin{bmatrix}\vec{x}\\ \vec{y}\end{bmatrix}_{i+1} = \begin{bmatrix}\vec{x}\\ \vec{y}\end{bmatrix}_{i} - s\nabla F = \begin{bmatrix}\vec{x}\\ \vec{y}\end{bmatrix}_{i} - s\begin{bmatrix}\frac{\partial F(\vec{x},\vec{y})}{\partial x_1}\\ \vdots\\ \frac{\partial F(\vec{x},\vec{y})}{\partial x_n}\\ \frac{\partial F(\vec{x},\vec{y})}{\partial y_1}\\ \vdots\\ \frac{\partial F(\vec{x},\vec{y})}{\partial y_n}\end{bmatrix}$$

Here $\nabla F$ is the gradient of F. For s we tried values ranging from 2^(1-h), where h ranged from 1 to 21, and hence s ranged from 1 to 9e-7. We selected the value of h that resulted in the lowest value of F. Next, we applied 20 iterations of Newton's method after gradient descent to quickly find local minima:

$$\begin{bmatrix}\vec{x}\\ \vec{y}\end{bmatrix}_{i+1} = \begin{bmatrix}\vec{x}\\ \vec{y}\end{bmatrix}_{i+1} - s \cdot J(F)^{-1} \cdot \nabla F = \begin{bmatrix}\vec{x}\\ \vec{y}\end{bmatrix}_{i+1} - s \cdot \begin{bmatrix}\frac{\partial^2 F(\vec{x},\vec{y})}{\partial^2 x_1} & \cdots & \frac{\partial^2 F(\vec{x},\vec{y})}{\partial x_1 \partial y_n}\\ \vdots & & \vdots\\ \frac{\partial^2 F(\vec{x},\vec{y})}{\partial x_n \partial y_1} & \cdots & \frac{\partial^2 F(\vec{x},\vec{y})}{\partial^2 y_n}\end{bmatrix}^{-1} \cdot \begin{bmatrix}\frac{\partial F(\vec{x},\vec{y})}{\partial x_1}\\ \vdots\\ \frac{\partial F(\vec{x},\vec{y})}{\partial x_n}\\ \frac{\partial F(\vec{x},\vec{y})}{\partial y_1}\\ \vdots\\ \frac{\partial F(\vec{x},\vec{y})}{\partial y_n}\end{bmatrix}$$

Here J(F) is the Jacobian of F. For s, we tried values ranging from 2^(1-h), where h ranged from 1 to 21. We selected the value of h that resulted in the lowest value of F. To calculate the gradient, we calculate the derivative of the objective function at x and y for each point:

$$\frac{\partial F(\vec{x},\vec{y})}{\partial x_k} = \sum_{i=1}^{n-1}\sum_{j=i+1}^{n} 2(\delta_{i=k} + \delta_{j=k}) w_{i,j} \frac{\left(\sqrt{(x_i - x_j)^2 + (y_i - y_j)^2} - d_{i,j}\right)}{\sqrt{(x_i - x_j)^2 + (y_i - y_j)^2}} (x_k - \delta_{i\neq k} x_i - \delta_{j\neq k} x_j)$$

$$\frac{\partial F(\vec{x},\vec{y})}{\partial y_k} = \sum_{i=1}^{n-1}\sum_{j=i+1}^{n} 2(\delta_{i=k} + \delta_{j=k}) w_{i,j} \frac{\left(\sqrt{(x_i - x_j)^2 + (y_i - y_j)^2} - d_{i,j}\right)}{\sqrt{(x_i - x_j)^2 + (y_i - y_j)^2}} (y_k - \delta_{i\neq k} y_i - \delta_{j\neq k} y_j)$$

Here $\delta_{i=j}$ is the Dirac delta function which is 1 if i and j are equal to each other, and 0 otherwise. Similarly, $\delta_{i\neq k}$ is 1 if i and k are equal to each other, and 0 otherwise. To calculate the Jacobian, we calculate each of the second derivatives empirically setting delta = 0.0000001:

$$\frac{\partial^2 F(\vec{x},\vec{y})}{\partial y_l \partial y_k} = \frac{\left.\frac{\partial F(\vec{x},\vec{y})}{\partial y_k}\right|_{yl+delta} + \left.\frac{\partial F(\vec{x},\vec{y})}{\partial y_k}\right|_{yl-delta}}{2 * delta}$$

We calculate other partial derivatives for all pairs *i* and *j* for both x and y coordinates similarly.

### Supplementary Figures

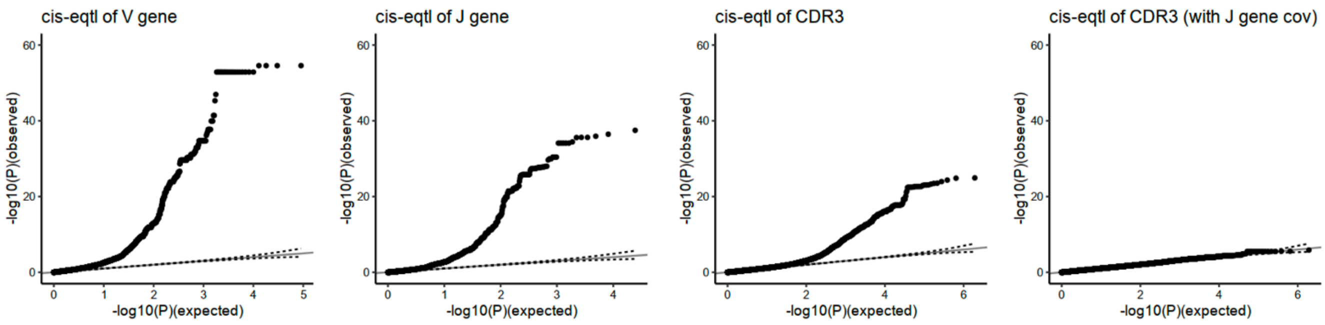


**Supplementary Figure 1. Cis-regulatory effects of CDR3 amino acid compositions.**

Using the replication dataset (n = 169), we tested associations between the variants within the *TCR* locus and V/J usage and CDR3 amino acid compositions. We also conducted the same analysis for CDR3 amino acid compositions but including covariates of nine J genes for which we observed significant cis-regulatory effects.

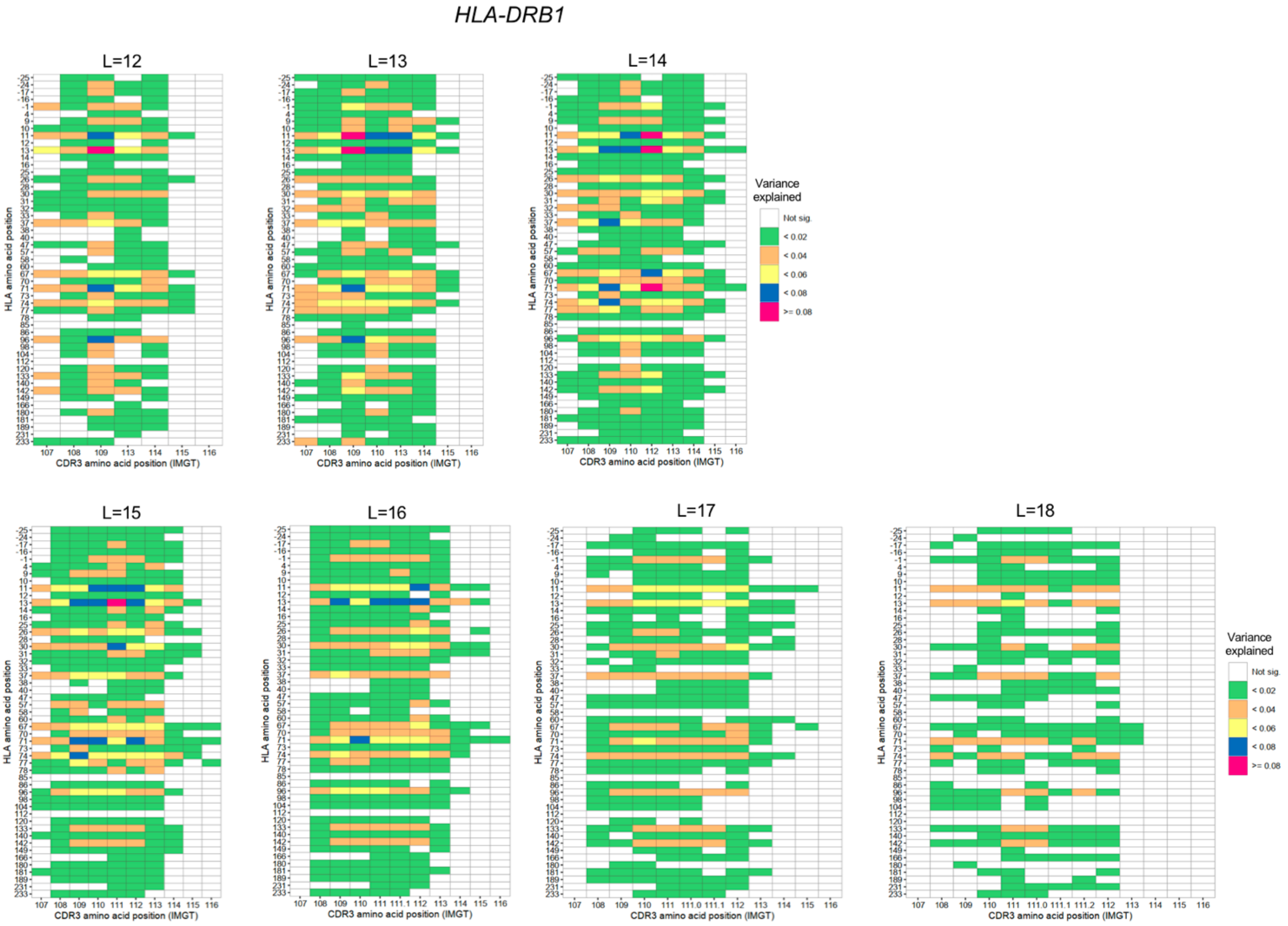


**Supplementary Figure 2. Variance explained in cdr3-QTL analysis for each length of CDR3.** Variance explained in cdr3-QTL analysis for different length of CDR3 (n=628; multivariate multiple linear regression). The results for *HLA-DRB1* were provided.

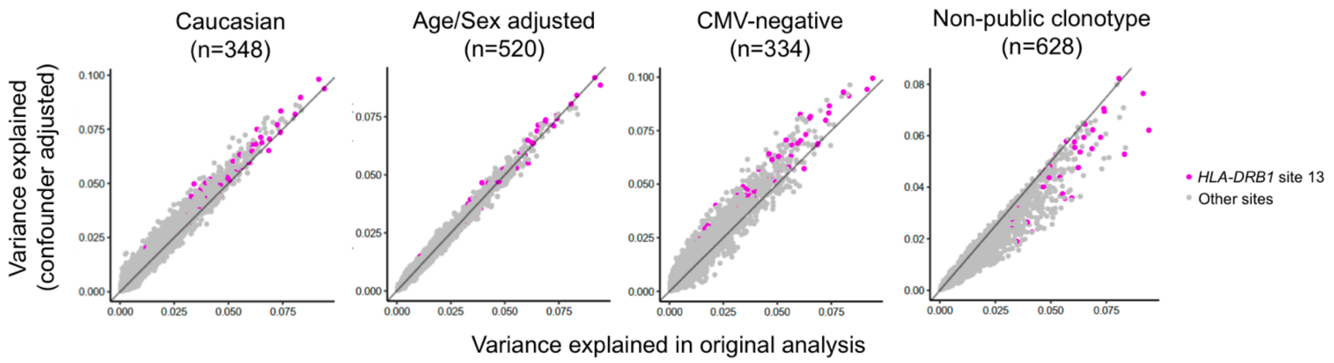


**Supplementary Figure 3. Evaluation of the confounders in cdr3-QTL analysis (multivariate multiple linear regression).**
To evaluate the effect of potential confounders in cdr3-QTL analysis, we tested variance explained using four different conditions (multivariate multiple linear regression). X-axis is the variance explained in the primary analysis (n=628) and Y-axis is the variance explained in each condition. We analyzed all tests in the primary analysis (24,430 tests). First, to test the potential bias by different ethnicities, we conducted the analysis only using Caucasian samples (n=348). Second, to test the potential bias by age and sex, we conducted the analysis by adding age and sex as covariates (n=520; sample size decreased due to missing data in covariates). Third, to test the potential bias by cytomegalovirus infection status, we conducted the analysis only using non-infected samples (n=334). Lastly, to test the potential bias by public clonotypes, we conducted the analysis excluding public clonotype (n=628).

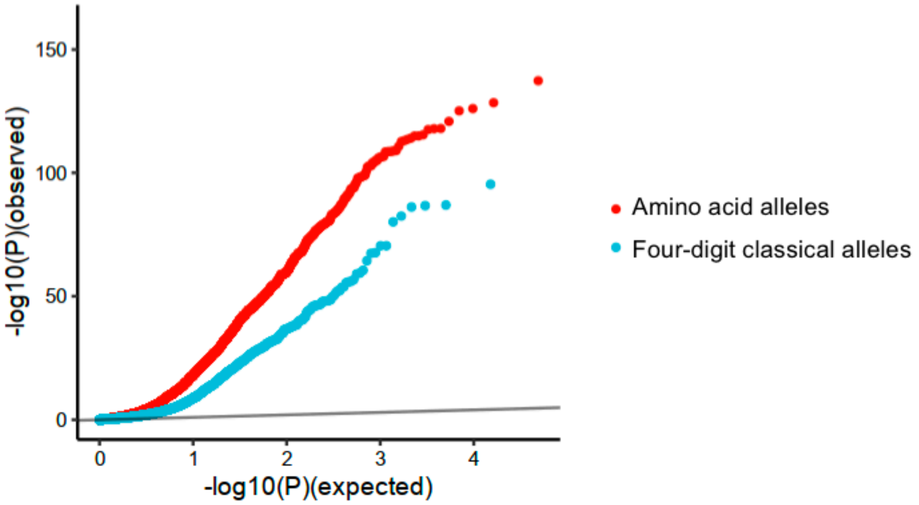


**Supplementary Figure 4. cdr3-QTL results using four-digit classical allele genotypes.**
QQ plot of cdr3-QTL analysis comparing two different methods (n = 628; multivariate multiple linear regression); associations with *HLA* amino acid alleles and those with four-digit classical *HLA* alleles.

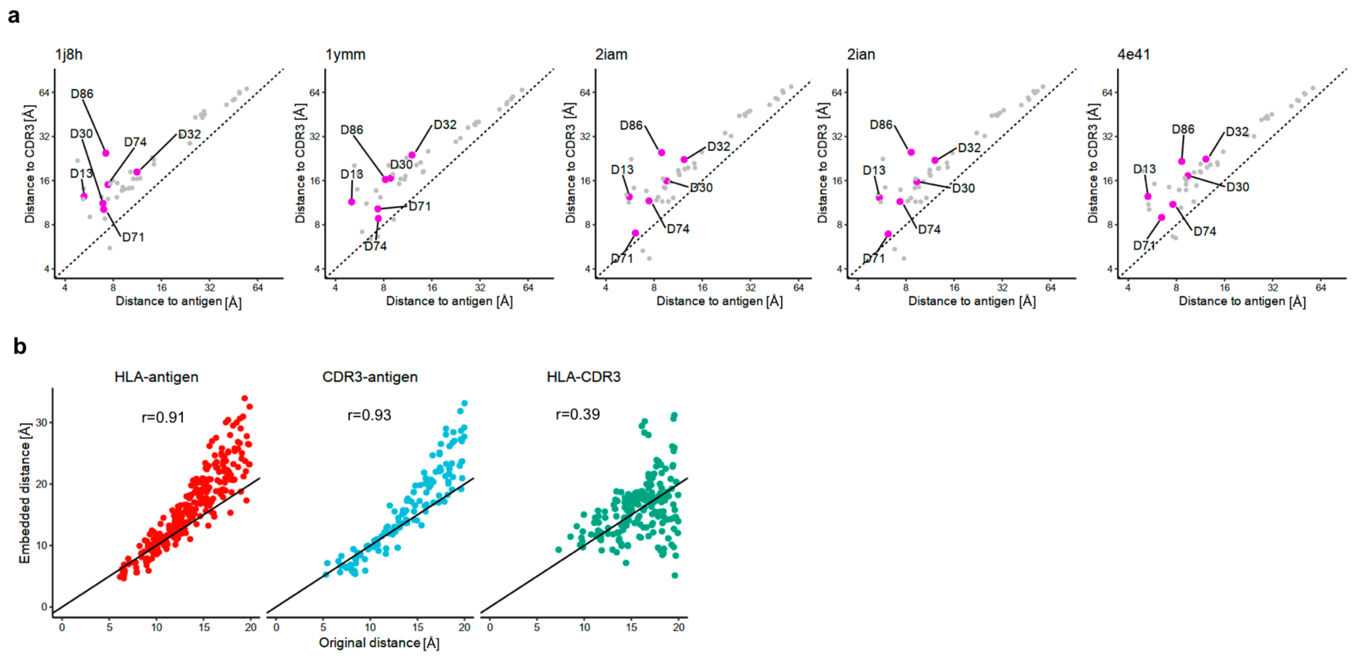


**Supplementary Figure 5. The observed and embedded pair-wise distances of amino acids in MHC-peptide-TCR complexes.**

**(a)** The shortest distances between each *HLA-DRB1* site and all positions of the antigen (X-axis) and those between each *HLA-DRB1* site and all positions of the CDR3 (Y-axis) were provided for each protein structure. The sites with independently significant cdr3-QTL effects were highlighted by magenta.

**(b)** Pair-wise distances in the original datasets and those in the embedded space. Averaged value across five structures were utilized. Two-dimensional embedding analysis based on the pair-wise distances was conducted (**Methods**). We down-weighted the distances between HLA and TCR so that their antigen-mediated indirect interaction was highlighted. Pearson's correlation coefficients were provided.

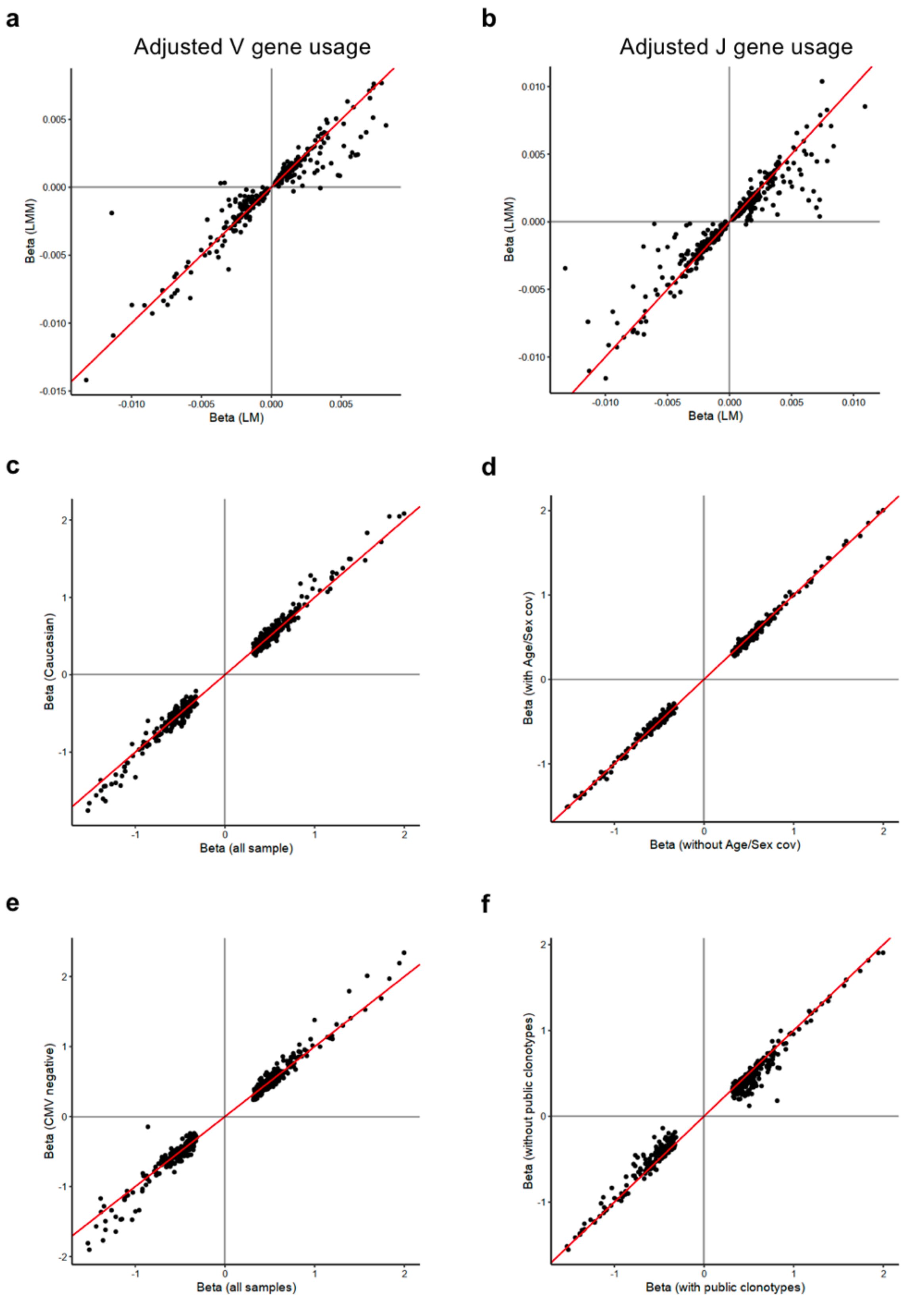


**Supplementary Figure 6. Evaluation of the confounders in cdr3-QTL analysis (liner regression).**
The analysis was restricted to the 435 CDR3 phenotypes (length-position-amino acid combinations) which had at least one significant association in the primary analysis (*P* < 0.05/1,262,664; linear regression).

**(a)** Effect sizes from a linear regression model where the effect of V genes was not adjusted (the primary analysis) and those from a linear mixed regression model where the effect of V genes was adjusted were compared. Effect sizes for non-normalized phenotype were used.
**(b)** Effect sizes from a linear regression model where the effect of J genes was not adjusted (the primary analysis) and those from a linear mixed regression model where the effect of J genes was adjusted were compared. Effect sizes for non-normalized phenotype were used.
**(c)** Effect sizes from a linear regression model where all samples were utilized (n=628; the primary analysis) and those where only Caucasian samples were utilized (n=348) were compared.
**(d)** Effect sizes from a linear regression model where the age and sex effects were not adjusted (n=628; the primary analysis) and those where the age and sex effects were adjusted (n=520) were compared. Since there were many missing values in covariate data, including them in a model decreased the sample size.
**(e)** Effect sizes from a linear regression model where all samples were utilized (n=628; the primary analysis) and those where only cytomegalovirus non-infected donors were utilized (n=334) were compared.
**(f)** Effect sizes from a linear regression model where all CDR3 were utilized (n=628; the primary analysis) and those where public clonotypes were excluded (n=628) were compared.

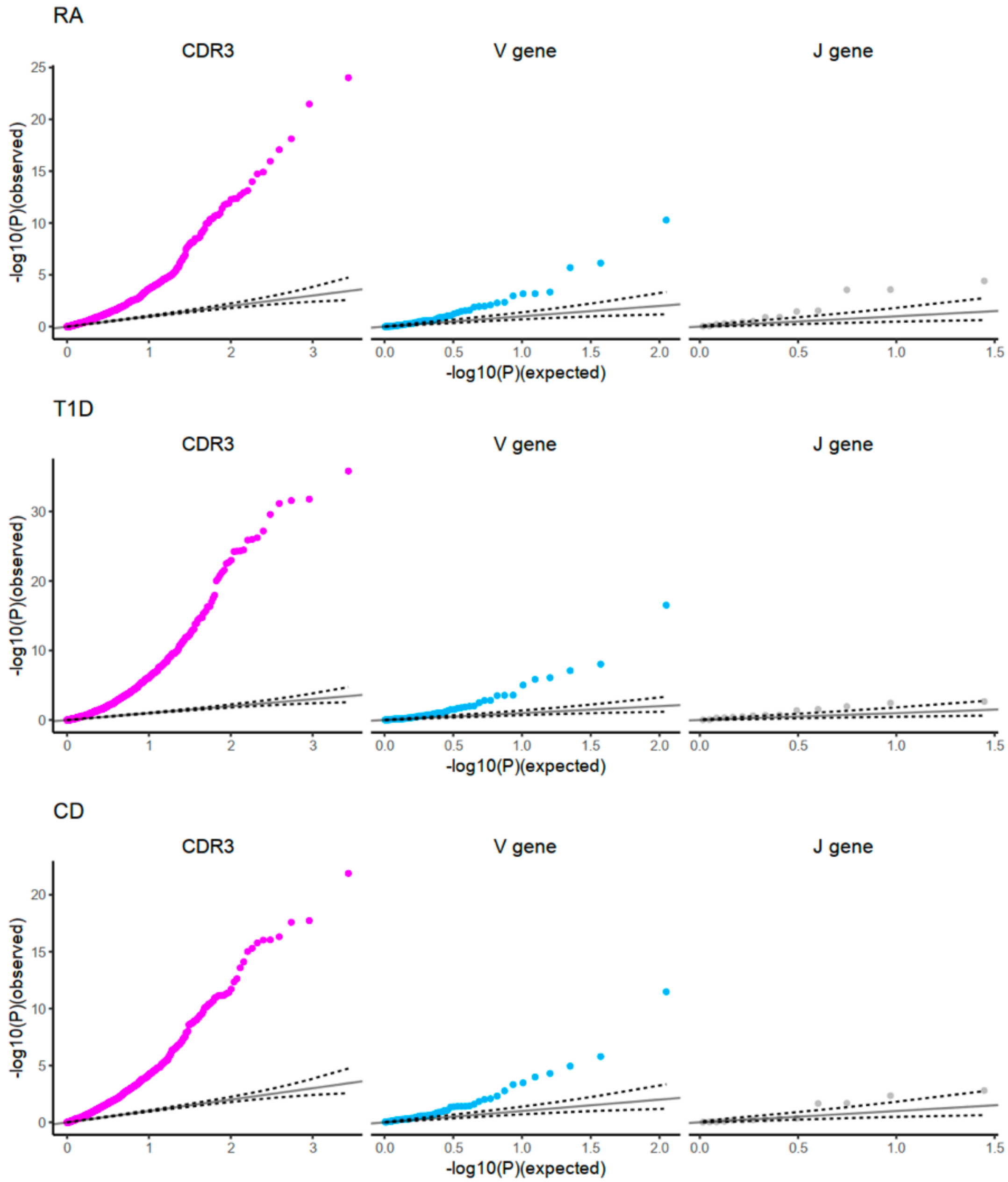


**Supplementary Figure 7. QQ plot of cdr3-QTL, V/J gene association based on *HLA*-risk scores.**
The identity line is provided with the 95% confidence interval.

**a**

*P* value threshold: 0.05

| Length | Pos. in CDR3 | Amino acids | Beta | *P* value |
|---|---|---|---|---|
| 15 | 110 | T | 0.19 | 0.0001 |
| ~~15~~ | ~~111~~ | ~~W~~ | ~~-0.15~~ | ~~0.1~~ |
| 15 | 113 | Q | 0.21 | 0.0003 |
| ⋮ | ⋮ | ⋮ | ⋮ | ⋮ |
| 15 | 115 | T | 0.1 | 0.001 |

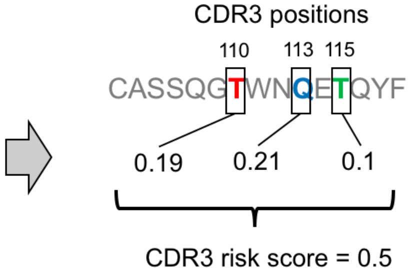


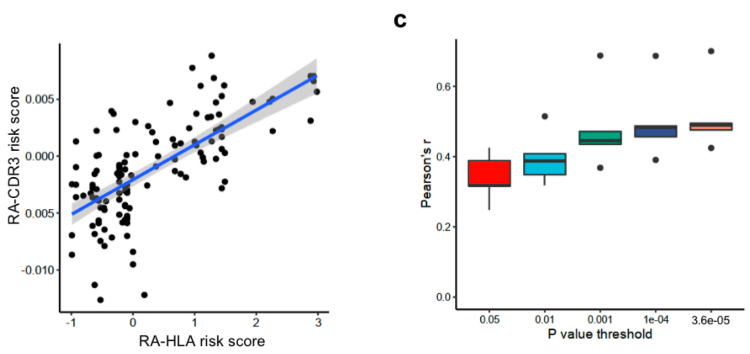


**Supplementary Figure 8. The strategy to calculate CDR3 risk score.**
**(a)** Schematic explanation of our strategy to calculate CDR3 risk score. The table shows the effect size estimate of cdr3-QTL analysis based on *HLA* risk scores. Effect sizes for corresponding amino acid positions are summed up to calculate the CDR3 risk score. Effect sizes which passed a *P* value threshold were utilized.
**(b)** Using the discovery dataset, we conducted 5-fold cross validation to evaluate the performance of RA-CDR3 risk score. A representative plot of RA-HLA risk score and RA-CDR3 risk score in a round of cross validation (with Bonferroni-corrected *P* < 0.05 threshold).
**(c)** The Pearson's correlation coefficient between RA-HLA risk score and RA-CDR3 risk score in the 5-fold cross validation using five different *P* value thresholds.
Within each boxplot, the horizontal lines reflect the median, the top and bottom of each box reflect the interquartile range (IQR), and the whiskers reflect the maximum and minimum values within each grouping no further than 1.5 x IQR from the hinge.
